# A susceptibility network analysis of disease trajectories leading to multiple sclerosis: a nationwide cohort study

**DOI:** 10.64898/2026.01.19.26344395

**Authors:** Ali Ebrahimi, Uffe Kock Wiil, Tomas Olsson, Pietro Lio, Ingrid Kockum, Ali Manouchehrinia, Narsis A. Kiani

## Abstract

**Background:** The prodromal phase of multiple sclerosis (MS) is increasingly recognized, but most studies have focused on isolated symptoms or static comorbidity counts, leaving the evolving structure of pre-onset disease burden underexplored.

**Objective:** To characterize dynamic disease trajectories preceding MS onset through longitudinal network modeling.

**Methods:** Health data from 10,273 MS patients and 47,167 matched controls in Sweden were analyzed. Disease co-occurrence networks were constructed for three pre-onset windows (0–5, 5–10, 10–15 years), with comparisons of centrality, clustering, and path length. Rewiring scores captured structural shifts, while Markov clustering and trajectory mapping identified comorbidity communities.

**Results:** MS networks were denser, more clustered, and showed shorter path lengths than controls, reflecting higher systemic interconnectivity. Psychiatric and metabolic diagnoses, especially depression, anxiety, diabetes, and abdominal pain, were hubs that gained prominence over time. Distinct clusters, including neuropsychiatric-toxicological and immune-endocrine constellations, were observed only in MS. Rewiring analysis revealed significant topological shifts in key diagnoses, such as inflammatory CNS disorders and substance use, as onset approached.

**Conclusions:** MS is preceded by dynamic reorganization of the comorbidity landscape, marked by increasing connectivity and rewired hubs. This framework highlights systemic disruption before diagnosis and provides a novel, network-based tool for studying prodromes in complex disorders.

## 1. Introduction

Multiple sclerosis (MS) is a chronic immune-mediated disorder of the central nervous system characterized by inflammation, demyelination, and neurodegeneration. Its etiology remains unresolved, with heritability, reflecting both genetic susceptibility and shared familial environmental factors, explaining ~50% of risk (1). Epstein–Barr virus (EBV) is now considered a near-obligate causal trigger, whereas other environmental modifiers such as tobacco smoke, low vitamin D, organic solvents, and adolescent obesity contribute additional risk (2). Increasing evidence supports a long prodromal phase, marked by greater healthcare use, psychiatric symptoms, infections, and musculoskeletal complaints years before diagnosis (3). In addition, conditions such as infectious mononucleosis (4), concussion (5, 6), thyroid disease (7), psoriasis (8), and inflammatory bowel disease (9) occur more frequently in MS, though most studies have examined only a limited set of comorbidities.

Nationwide health registries now allow systematic assessment of disease associations and their combined contribution to MS risk. Network science offers a framework for mapping these associations, characterizing diseases as nodes and co-occurrences as edges. Several studies have previously utilized network approaches to understand disease associations and comorbidities (10–12). While network approaches are established in molecular biology and brain connectivity research, their application at the population level to study MS phenotypes remains limited. Importantly, most prior studies provide static snapshots of comorbidities rather than examining how networks evolve during the prodrome.

Dynamic comorbidity network analysis can reveal how stable diagnostic constellations and rewired interactions contribute to latent disease risk. Identifying key hub conditions and their temporal organization may support earlier recognition of MS.

Here, we performed a large-scale registry-based study of individuals with MS and matched controls, constructing weighted longitudinal comorbidity networks across three pre-onset windows and applying graph-theoretical, clustering, and rewiring analyses to characterize the structure and temporal evolution of the MS prodrome.

## 2. Methods

### 2.1. Data source

Diagnoses were obtained from the Swedish National Patient Register (NPR) (13), which includes nationwide inpatient data since 1987 and hospital-based outpatient specialist visits since 2001, but does not capture primary care. Accordingly, disease histories reflect temporal sequences of ICD-10 diagnoses recorded in specialist care rather than detailed clinical case histories. The NPR has demonstrated high validity for chronic and neurological conditions, including MS (13–15).

MS cases were identified using the Swedish MS Registry (SMSreg) (15, 16) and/or the NPR. Individuals were classified as MS if registered in SMSreg or if they had ≥3 MS diagnoses (ICD-10: G35) in the NPR, a validated repeated-code definition that reduces misclassification from preliminary assessments (13). SMSreg provides clinician-confirmed diagnoses with longitudinal follow-up and covers approximately 85–90% of the Swedish MS population (15, 16), while the NPR ensures complete national coverage of inpatient and specialist outpatient care across all medical disciplines. Using both sources maximized diagnostic specificity and temporal completeness of pre-onset comorbidities (15).

For each MS case, five population-based controls matched on age, sex, index year, and residential area were selected. The MS index date was defined as the earliest of recorded onset in SMSreg, first MS diagnosis, or first demyelinating diagnosis (G36.0, G36.8–G36.9, G37.8–G37.9) in the NPR; controls were assigned the corresponding index date (17, 18).

To construct disease co-occurrence networks, individuals were required to have ≥2 outpatient ICD codes recorded before the index date, ensuring definable diagnostic transitions. Index dates were restricted to after January 1, 2005 to guarantee at least five years of outpatient NPR coverage for all participants.

### 2.2. Disease Trajectory Formation

We began by constructing a sequential disease trajectory for each individual, in which every unique ICD-10 code was treated as a separate diagnosis event (i.e., disease A → disease B → disease C → …). A diagnosis was counted as present if it occurred at least once before the index date. Even a single code was considered informative, as it reflects that the diagnosis was at least under clinical consideration. This choice prioritises sensitivity to capture diagnostic trajectories for network construction rather than strict confirmation of individual diseases.

Weighted, directed disease networks were constructed separately for MS and control groups by aggregating individual diagnostic trajectories. Nodes represent unique ICD-10 diagnoses, and directed edges indicate temporally ordered transitions between adjacent diagnoses within a patient’s history. No time window was imposed, preserving longitudinal progression rather than same-visit co-occurrence. To ensure network stability, only transitions observed in >10 individuals were retained, reducing noise from rare or idiosyncratic patterns. Edge weights reflected the number of patients exhibiting each transition.

### 2.3. Graph-Theoretical Analysis

We quantified global structural features of the largest weakly connected component in each network to compare breadth, connectivity, clustering, and community structure. Connectivity refers to the number of direct links each diagnosis has to other diagnoses, while breadth reflects the diversity of diagnostic categories it connects to. The global clustering coefficient measures the extent to which diagnoses linked to the same diagnosis are also linked to each other, indicating tightly interrelated diagnostic patterns. Community structure (network modularity) captures the presence of diagnostic groups that co-occur more frequently with one another than with the rest of the network.

We assessed community structure using modularity, which quantifies how strongly the network divides into densely connected groups. Communities were detected with the Louvain algorithm, which iteratively maximizes modularity.

To assess diagnostic influence, we applied complementary centrality measures: degree centrality (DC; number/strength of connections), closeness centrality (CC; distance to other diagnoses), betweenness centrality (BC; frequency on shortest paths), PageRank (PR; influence through links to other influential diagnoses), and local transitivity (clustering among a diagnosis’s neighbors).

To reduce sample-size bias, controls were down-sampled to the MS cohort size across 20 resampling rounds. For each resample, weighted, directed networks were reconstructed and centrality metrics recalculated on the largest component. Centrality distributions were compared using Wilcoxon tests on log-transformed values, and ranking stability was assessed by top-10 consistency across resamples.

### 2.4. Network clustering

To identify densely connected subgroups of diseases, we applied the Markov Cluster Algorithm (MCL) (19) using the clusterMaker2 app in Cytoscape with an inflation parameter of 3.0, which increases cluster granularity and has been used in prior clinical network studies. Prior to clustering, the networks were weighted, self-loops were removed, and the algorithm was applied to the largest weakly connected component. MCL partitions networks into non-overlapping communities by simulating flow dynamics, such that nodes within a cluster are more strongly connected to each other than to nodes outside the cluster. All other MCL settings were kept at their default clusterMaker2 parameters.

For MS and control networks, clusters were ranked by size and the 10 largest were retained. Clustered networks were exported to R as igraph objects, including only nodes assigned to MCL clusters 1–10. The dominant ICD chapter within each cluster was defined by the most frequent first-character ICD-10 code.

Cluster-level metrics included node and edge counts, density, average degree, node strength (weighted degree), and heterogeneity measured by the number of ICD chapters and the Shannon diversity index. The Shannon Index was selected because it captures the richness and distribution of diagnoses within a cluster and is increasingly used in multimorbidity research. It was calculated as *H* = − ∑ *p*^*i*^*ln*(*p*^*i*^), where *p*^*i*^ is the proportion of diagnostic events belonging to diagnosis *i*. Differences of approximately ≥0.10 or exceeding the 95% CI of controls were considered clinically meaningful. Clusters were clinically anchored by reporting the dominant ICD chapter and the top five diagnoses ranked by strength and degree, summarized into short descriptive labels.

### 2.5. Network rewiring

Network rewiring analysis was applied to identify (1) diagnoses showing the largest changes in connectivity within MS patients across three pre-onset windows (15–10, 10–5, and 5–0 years before onset), representing early, mid, and late prodromal periods, and (2) diagnoses exhibiting different network roles in MS compared with controls. Rewiring scores were calculated using the DyNet framework in Cytoscape, with diagnoses aggregated to the two-character ICD level (e.g., F32 “Depressive episode” grouped as F3 “Mood disorders”).

Rewiring scores quantified how much a diagnosis’ connectivity pattern changed under random network perturbation. Scores were z-standardized within each network, and values above the 90th percentile were defined a priori as highly rewired, capturing diagnoses with structurally unstable network positions. High scores indicate roles dependent on specific connections, whereas low scores reflect stable, resilient positions.

All statistical analyses and data management were performed in R version 4.2.1. Network clustering and dynamic analyses were conducted in Cytoscape version 3.9.1 using the MCL and DyNet algorithms (20, 21).

## 3. Results

From the initial 10,989 cases and 52,850 controls, we excluded 1,106 cases and 8,003 controls who had no pre-index ICD records. 193 cases with pre-index demyelinating codes recorded only as secondary diagnoses were removed to maintain a consistent and accurate case definition. Finally, 900 cases and 5,850 controls with fewer than two pre-index ICD codes were excluded to ensure sufficient data for network construction. The final analytic cohort included 8,790 cases (68.7% from the SMSreg and 31.3% from the NPR) and 38,997 controls. Characteristics of individuals with and without MS are shown in Table 1; groups were well matched with no significant differences in demographic or clinical characteristics.

**Table 1.** General demographic and clinical characteristics of the study population.

|  | MS (N=8,790) | Control (N=38,997) |
| --- | --- | --- |
| **Sex (%)** |  |  |
| Female | 6189 (70.4%) | 27684 (71.0%) |
| Male | 2601 (29.6%) | 11313 (29.0%) |
| **Age at Index** |  |  |
| Mean (SD) | 40.9 (14.1) | 40.7 (14.5) |
| Median (Q1, Q3) | 39.5 (29.8, 50.6) | 39.1 (29.2, 50.6) |
| **Calendar year of birth** |  |  |
| Mean (SD) | 1971 (15.0) | 1971(15.3) |
| Median (Q1, Q3) | 1972 (1961, 1983) | 1972 (1961, 1983) |
| **Age at first recorded ICD code** |  |  |
| Mean (SD) | 32.5 (14.9) | 32.2 (15.2) |
| Median (Q1, Q3) | 31 (21, 43) | 31 (21, 42) |
| **Age at last recorded ICD code** |  |  |
| Mean (SD) | 39.5 (14.5) | 38.7 (14.8) |
| Median (Q1, Q3) | 38 (28, 50) | 37 (27, 49) |
| **Average number of ICD codes** |  |  |
| Mean (SD) | 15.1 (19.6) | 14.2 (21.7) |
| Median (Q1, Q3) | 9 (4, 17) | 8.0 (4, 16) |
| **Follow-up** |  |  |
| Mean (SD) | 11 (3.9) | 11.1 (3.7) |
| Median (Q1, Q3) | 10.7 (7.6, 14.2) | 10.9 (7.9, 14.1) |
| **Index year** |  |  |
| 2001-2010 | 3615 (41.1%) | 16385 (42.0%) |
| 2011-2020 | 5175 (58.9%) | 22612 (58.0%) |
| **Socioeconomic status (quantiles)** |  |  |
| 1 (least affluent) | 1253 (14.3%) | 8161 (20.9%) |
| 5 | 1594 (18.1%) | 9199 (23.6%) |
| 3 | 2296 (26.1%) | 6911 (17.7%) |
| 2 | 1651 (18.8%) | 9418 (24.2%) |
| 4 (most affluent) | 1996 (22.7%) | 5308 (13.6%) |
| **Area of residence** |  |  |
| Rural | 2032 (23.1%) | 8983 (23.0%) |
| Urban | 6758 (76.9%) | 30014 (77.0%) |

2 SD: Standard deviation, ICD: International Classification of Diseases, Q: quartile

### 3.1. Structural Differences Between MS and Control Disease Networks

#### Overall Network Topology

Figure 1 shows the MS and control comorbidity networks restricted to the 50 highest in-degree diagnoses. The MS network contained fewer nodes (298 vs. 602) and exhibited lower density (0.011 vs. 0.013), average degree (6.33 vs. 15.30), and reciprocity (0.75 vs. 0.80), indicating more concentrated and less recurrent diagnostic transitions. MS networks showed longer average path lengths (3.04 vs. 2.71) but similar diameter (7 vs. 8) and clustering coefficients (0.201 vs. 0.208). Degree assortativity was less negative in MS (−0.02 vs. −0.18), suggesting greater hub–hub connectivity, while modularity was substantially higher (0.70 vs. 0.50), indicating more distinct diagnostic communities (Table S1).

**Figure 1.**
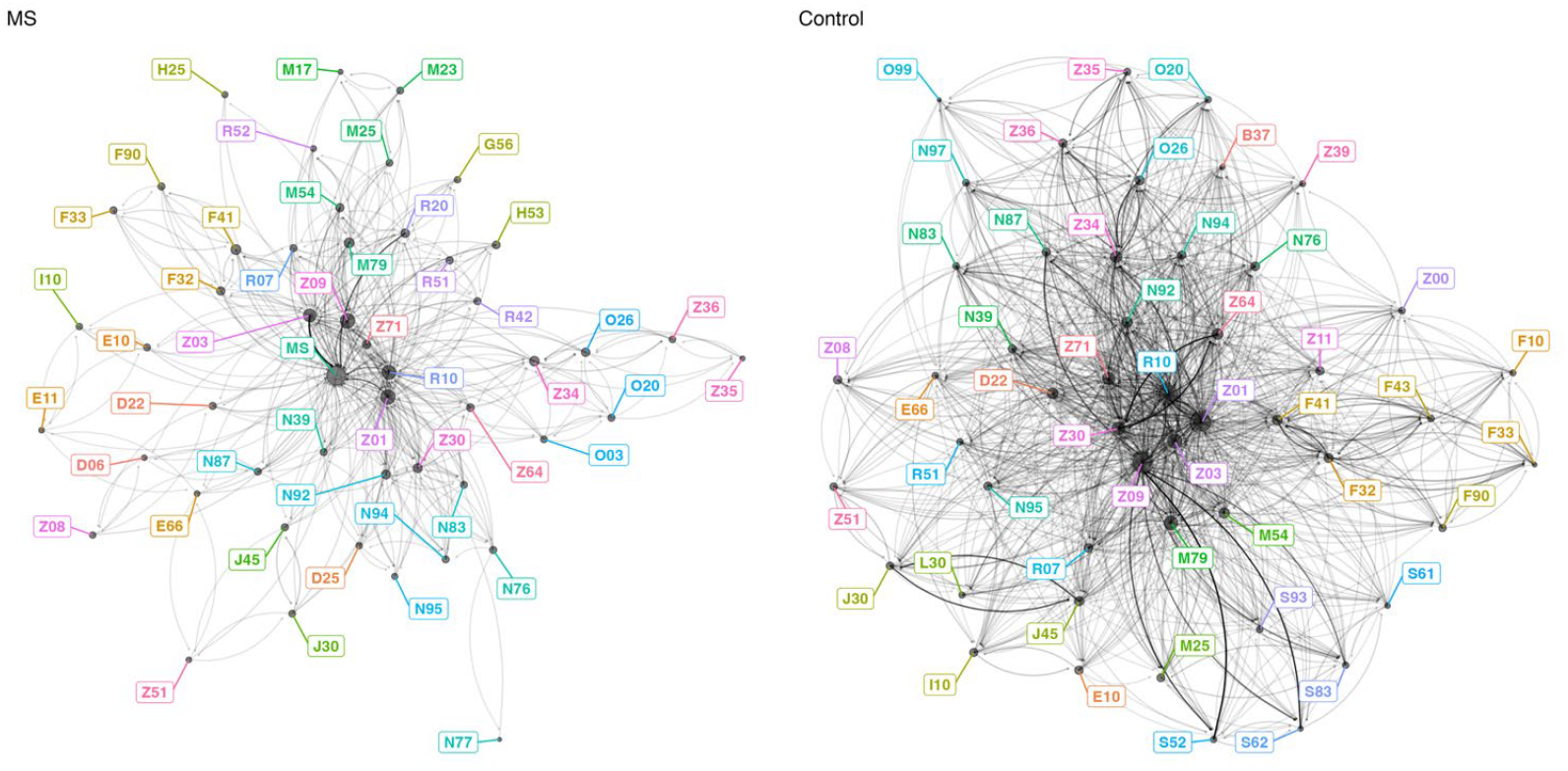
Top 50 most connected diagnoses in the multiple sclerosis (MS; left) and control (right) comorbidity networks. Nodes represent three-character ICD-10 codes, colored by ICD chapter, and sized by degree centrality (number of direct connections). Edges represent co-occurrence of diagnoses within individuals, with arrow direction indicating the temporal sequence of diagnoses and edge transparency proportional to the number of patients sharing that link. Only the 50 nodes with the highest in-degree are displayed for each network to emphasize the most prominent comorbidity patterns. ICD-10 chapter ranges shown in the figure correspond to the following categories: A00–B99 (infectious/parasitic), C00–D48 (neoplasms), D50–D89 (blood/immune), E00–E90 (endocrine/metabolic), F00–F99 (mental/behavioural), G00–G99 (nervous system), H00–H59 (eye), H60–H95 (ear/mastoid), I00–I99 (circulatory), J00–J99 (respiratory), K00–K93 (digestive), L00–L99 (skin), M00–M99 (musculoskeletal), N00–N99 (genitourinary), O00–O99 (pregnancy/childbirth), P00–P96 (perinatal), Q00–Q99 (congenital), R00–R99 (symptoms/signs), S00–T98 (injury/poisoning), V01–Y98 (external causes), and Z00–Z99 (health-status factors).

All node-level centrality measures differed significantly between MS and control networks (Figure 2), although effect sizes were small. MS diagnoses showed higher degree and strength, indicating more frequent and broader diagnostic co-occurrence, and higher closeness, reflecting more central positioning and shorter diagnostic paths. Local transitivity and PageRank were also modestly elevated, suggesting tighter clustering and greater overall influence of MS-related diagnoses. In contrast, betweenness was slightly lower, indicating that MS diagnoses less often functioned as bridges between otherwise weakly connected conditions.

**Figure 2.**
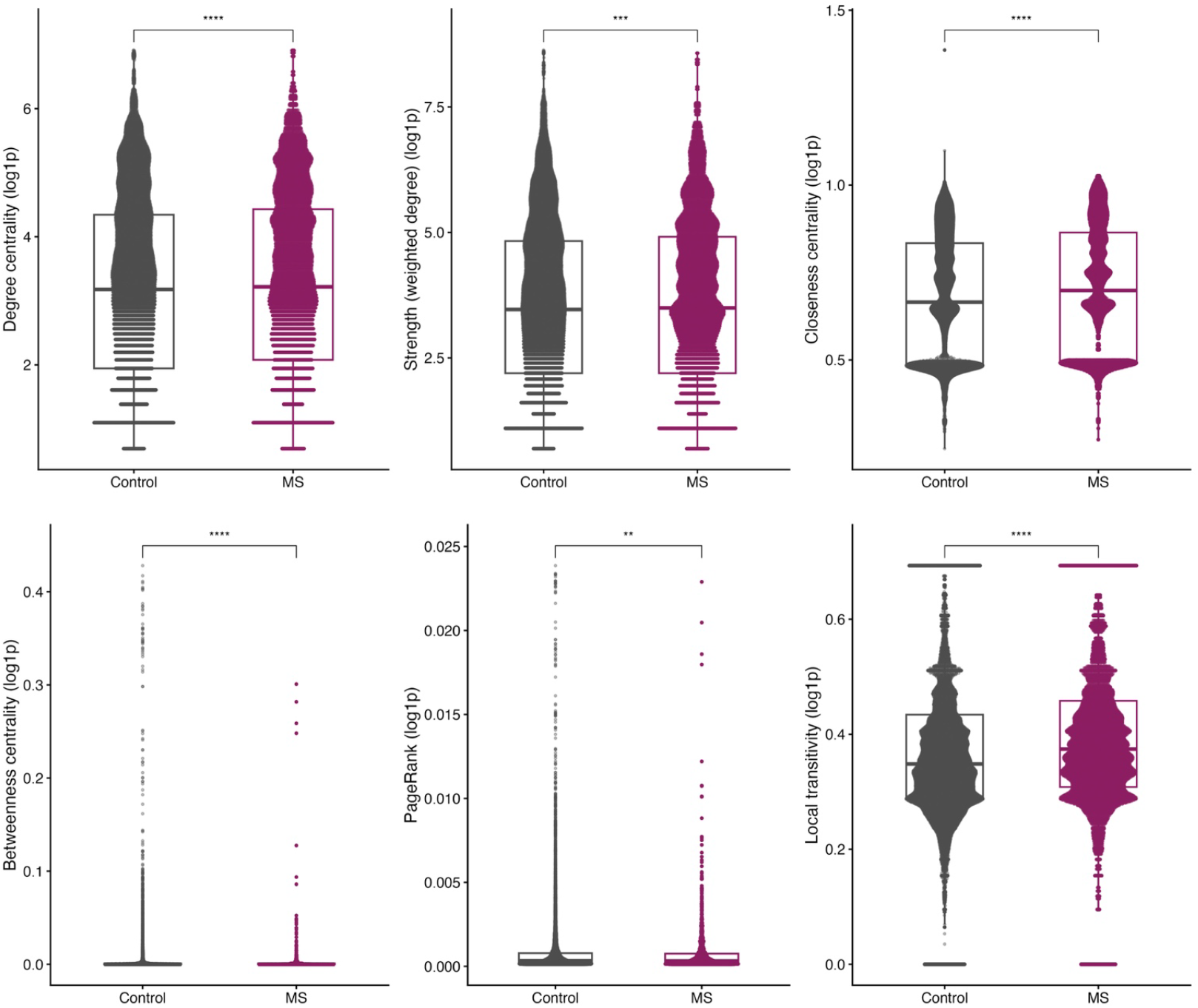
Comparison of node-level centrality measures between MS and control networks. Violin plots show the distribution of log-transformed centrality values for all nodes across both groups.

### 3.2. Central Nodes and Diagnostic Influence

With Z-codes and R10 included, both MS and control networks were dominated by non-specific codes (Z01, Z09, R10). After their exclusion, both networks were characterized by shared high-centrality diagnoses, including anxiety (F41), depression (F32), soft tissue pain (M79), and type 1 diabetes (E10). This overlap suggests that network structure is driven primarily by common comorbidities and healthcare utilization rather than MS-specific conditions, with only minor MS-related differences (e.g., R20 sensory disturbances). Thus, MS prodromal differentiation arises from the configuration of common diagnoses rather than rare or unique ones. Table S2 reports the top-ranked diseases across centrality measures (DC, CC, BC, EC, PR) for both groups.

### 3.3. Clustering Reveals Diagnostic Structures Unique to MS

Clear differences were observed among the top 10 clusters in MS and control networks. MS clusters were dominated by mental and behavioural disorders, endocrine/metabolic conditions, respiratory diseases, pregnancy-related complications, and general symptoms, whereas control clusters more often centered on infectious, neurologic, ophthalmologic, dermatologic, genitourinary, and congenital conditions; digestive diseases and injuries appeared in both groups. MS clusters were more multisystem, spanning a higher number of ICD chapters on average (12.6 vs. 7.7) and showing greater heterogeneity (higher Shannon index), indicating broader comorbidity patterns than the more siloed control clusters (Table 2).

**Table 2:** Dominant clinical clusters (top 10) in MS and control networks based on ICD-coded disease co-occurrence patterns. Each cluster represents a group of frequently co-occurring conditions. The dominant ICD chapter indicates the most frequent diagnostic category within the cluster. Anchor conditions are the top-ranking ICD codes contributing to each cluster’s structure and clinical theme

|  | Cluster | Dominant ICD chapter | Top anchor conditions | Total number of ICDs | Shannon index |
| --- | --- | --- | --- | --- | --- |
| MS | 1 | R – Symptoms/signs | Dizziness/giddiness; Headache; Chest/throat pain; Dyspnea; Palpitations | 142 | 2.76 |
|  | 2 | S – Injury/poisoning | Fracture of lower leg; Fracture of forearm; Fracture of foot; Joint sprain; Shoulder/upper arm fracture | 106 | 2.41 |
|  | 3 | K – Digestive | Cholelithiasis; GERD; IBS; Hemorrhoids; Diverticular disease | 89 | 2.48 |
|  | 4 | E – Endocrine/metabolic | Type 1 diabetes; Type 2 diabetes; Unspecified diabetes; Hypothyroidism; Dyslipidemia | 45 | 2.58 |
|  | 5 | C – Neoplasms | Breast cancer; Corpus uteri cancer; Testis cancer; Nodal metastasis | 42 | 1.91 |
|  | 6 | F – Mental/behavioral | Anxiety disorders; Personality disorders; Alcohol-related disorders | 35 | 1.82 |
|  | 7 | O – Pregnancy/childbirth | Antenatal/obstetric conditions; Labour complications; Obstetric trauma | 30 | 0.96 |
|  | 8 | F – Mental/behavioral | Depressive episode; Somatoform disorders; Sleep disorders; Anxiety; Adjustment disorders | 25 | 2.18 |
|  | 9 | F – Mental/behavioral | Hyperkinetic disorders; Alcohol-related disorders; Other substance use; Personality disorders | 20 | 1.54 |
|  | 10 | J – Respiratory | Asthma; Allergic rhinitis; Pneumonia; Chronic sinusitis | 17 | 1.61 |
| **Controls** | 1 | S – Injury/poisoning | Knee sprain; Forearm fracture; Wrist/hand fracture; Lower leg fracture; Open wound of wrist/hand | 808 | 2.97 |
|  | 2 | Q – Congenital | Phakomatoses; Urinary tract malformations; Cardiovascular malformations; Chromosomal deletions/abnormalities | 29 | 2.18 |
|  | 3 | L – Skin | Acne; Seborrhoeic keratosis; Pigment disorders; Dermatitis; Psoriasis | 14 | 1.67 |
|  | 4 | H – Eye/ear | Refraction disorders; Strabismus; Conjunctival disorders; Glaucoma; Retinal disease | 13 | 1.31 |
|  | 5 | N – Genitourinary | Chronic kidney disease; Obstructive/reflux uropathy; Cystitis; UTI; Urolithiasis | 12 | 0.84 |
|  | 6 | G – Nervous system | Polyneuropathy; Cerebrovascular syndromes; Parkinson's disease; Epilepsy | 11 | 2.02 |
|  | 7 | H – Eye/ear | Keratitis; Lacrimal disorders; Scleral inflammation; Glaucoma | 10 | 1.28 |
|  | 8 | L – Skin | Dermatitis; Allergic contact dermatitis; Lichen simplex chronicus; Urticaria; Psoriasis | 10 | 1.09 |
|  | 9 | K – Digestive | Ulcerative colitis; Noninfective gastroenteritis; IBS; Cholelithiasis; Diverticular disease | 9 | 1.68 |
|  | 10 | B – Infectious | Chronic viral hepatitis; HIV; Syphilis; Tuberculosis | 8 | 1.39 |

Several clinical themes were unique to MS clusters, including psychiatric and substance use disorders, mood disorders, metabolic–endocrine conditions (e.g., hypothyroidism, diabetes, dyslipidemia), and certain solid tumors. Psychiatric diagnoses did not form a single cluster: mood disorders (F3) clustered separately from anxiety, personality, substance-related, developmental, and behavioural disorders (Clusters 6, 8, and 9). In contrast, control-only clusters were driven by chronic viral infections, ophthalmologic disorders, inflammatory polyneuropathies, and cerebrovascular syndromes. Together, these patterns reinforce that the MS prodrome involves multisystem convergence rather than single-specialty comorbidity. Supplementary Tables S3–S8 and Figures S1–S2 provide cluster details.

### 3.4. Rewiring Analysis Highlights Shifts in Disease Connectivity in MS

In MS–control comparisons (Fig. 3), the most rewired diagnoses spanned immune/inflammatory and infectious categories (M0, M3, E8, J1), neurologic and neuropsychiatric presentations (G9, H5, R4), cutaneous/sensory symptoms (R2), and systemic risk factors (Z9, N1). Sensory disturbances (R2), including paresthesias (R20), showed the strongest rewiring, with high PageRank and betweenness, indicating a hub role linking psychiatric, immune/infectious, and metabolic domains. While local connectivity generally decreased, betweenness increased in MS, particularly for R2, G9, and R4, and several nodes (R2, E8, G9) gained global influence despite fewer connections, consistent with fewer but more consequential transitions.

**Figure 3.**
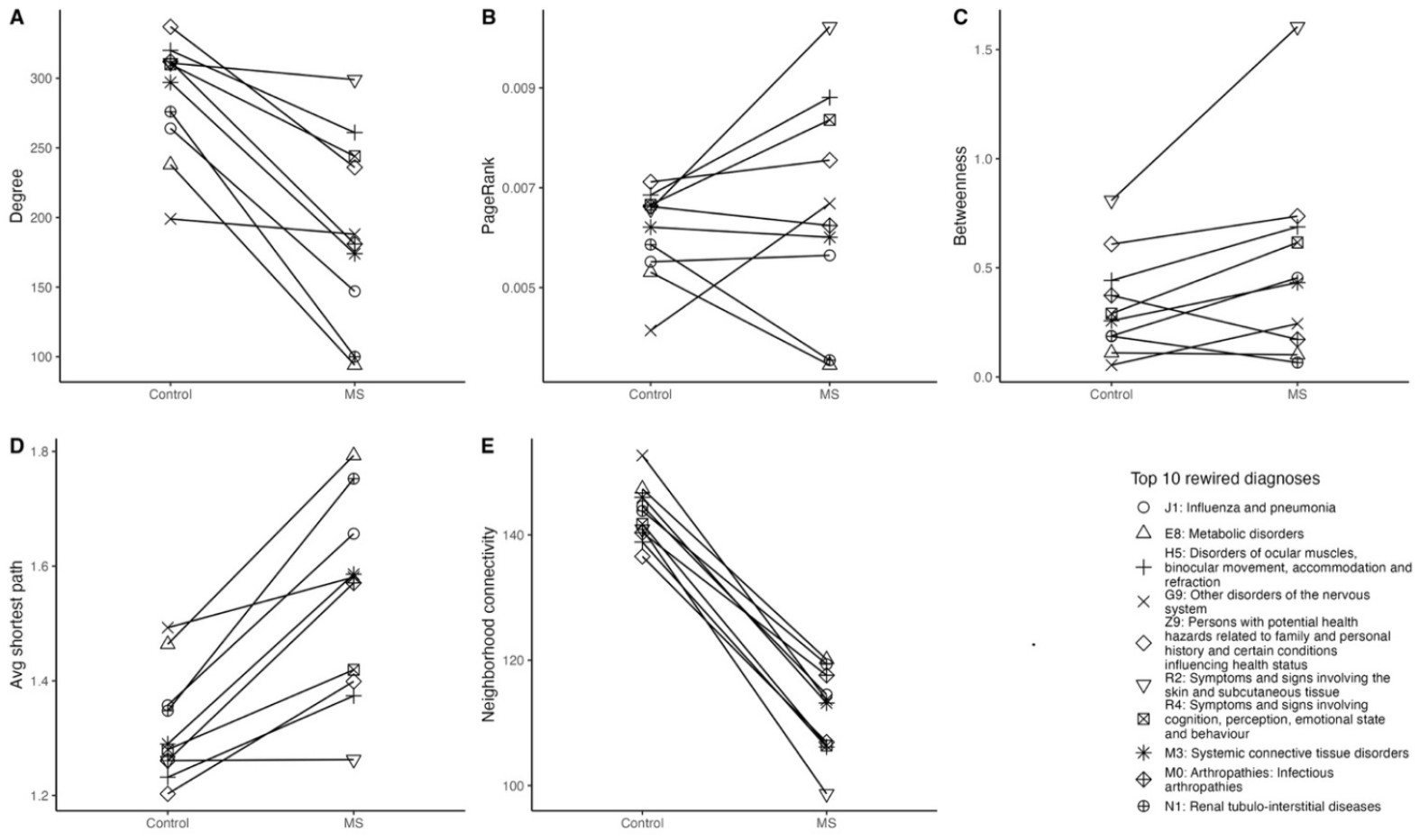
Comparison of centrality for the top 10 rewired diagnoses selected by DyNet score between MS and controls.

**Figure 4.**
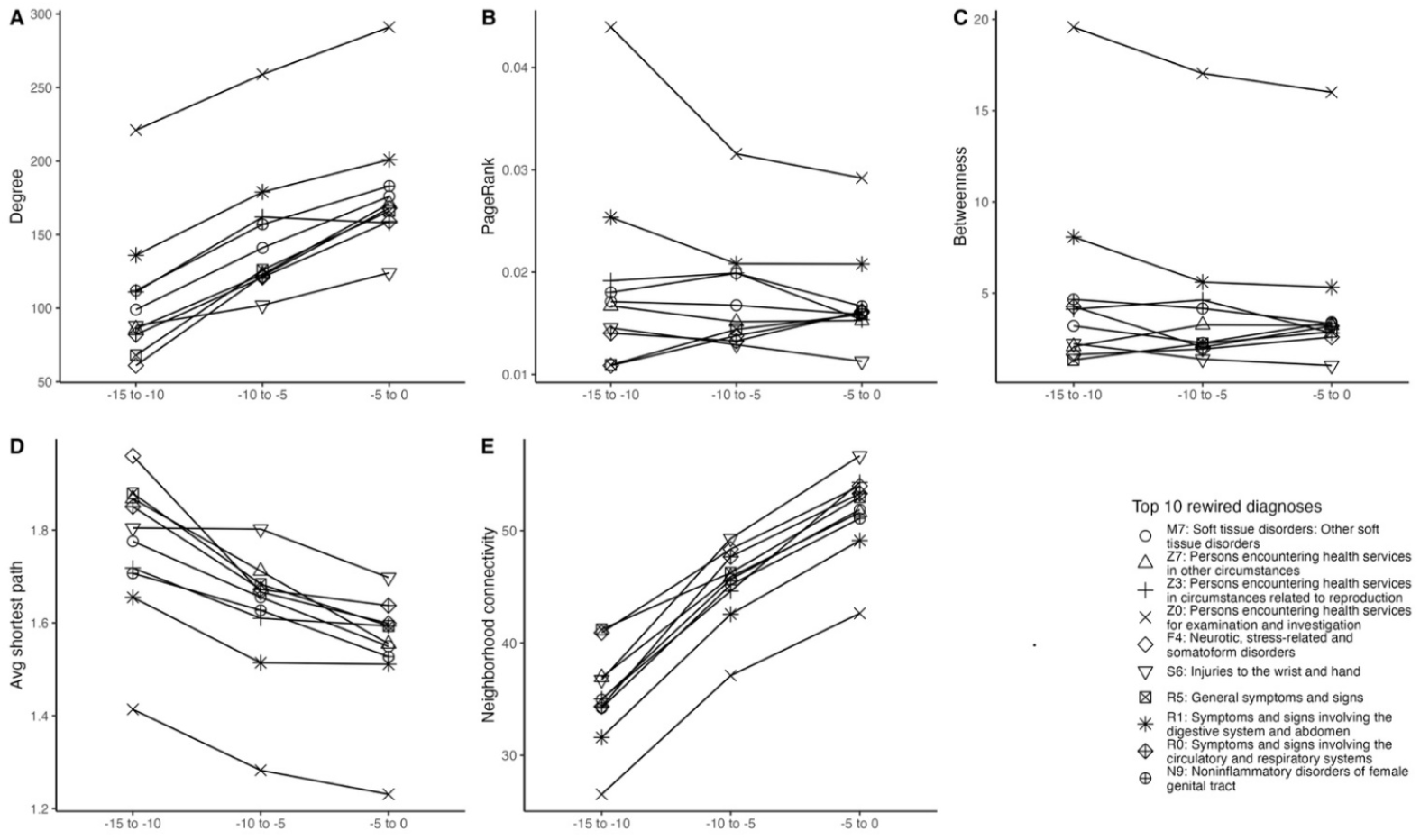
Comparison of centrality for the top 10 rewired diagnoses selected by DyNet score in MS cases over the past 15 years before MS onset.

Within the MS cohort across the 15-year pre-onset period, the most rewired diagnoses spanned musculoskeletal conditions (M7), health-status and healthcare-contact codes (Z0, Z3, Z7), mental and behavioural disorders (F4), injury-related conditions (S6), general symptoms and abnormal findings (R0, R1, R5), and genitourinary disorders (N9). These nodes showed increasing connectivity and shorter path lengths toward onset, indicating late-prodromal convergence. General examination (Z0) emerged as the primary hub or bridge, with neurotic and stress-related disorders (F4) gaining influence as secondary connectors, while symptom-based (R0/R1/R5), healthcare-contact (Z3/Z7), musculoskeletal (M7), and injury-related (S6) codes became progressively integrated into the network.

In matched controls over the same 15-year period (Fig. 5), rewiring was uniform and lacked centrality convergence, supporting MS-specific rather than general healthcare effects (Table S9).

**Figure 5.**
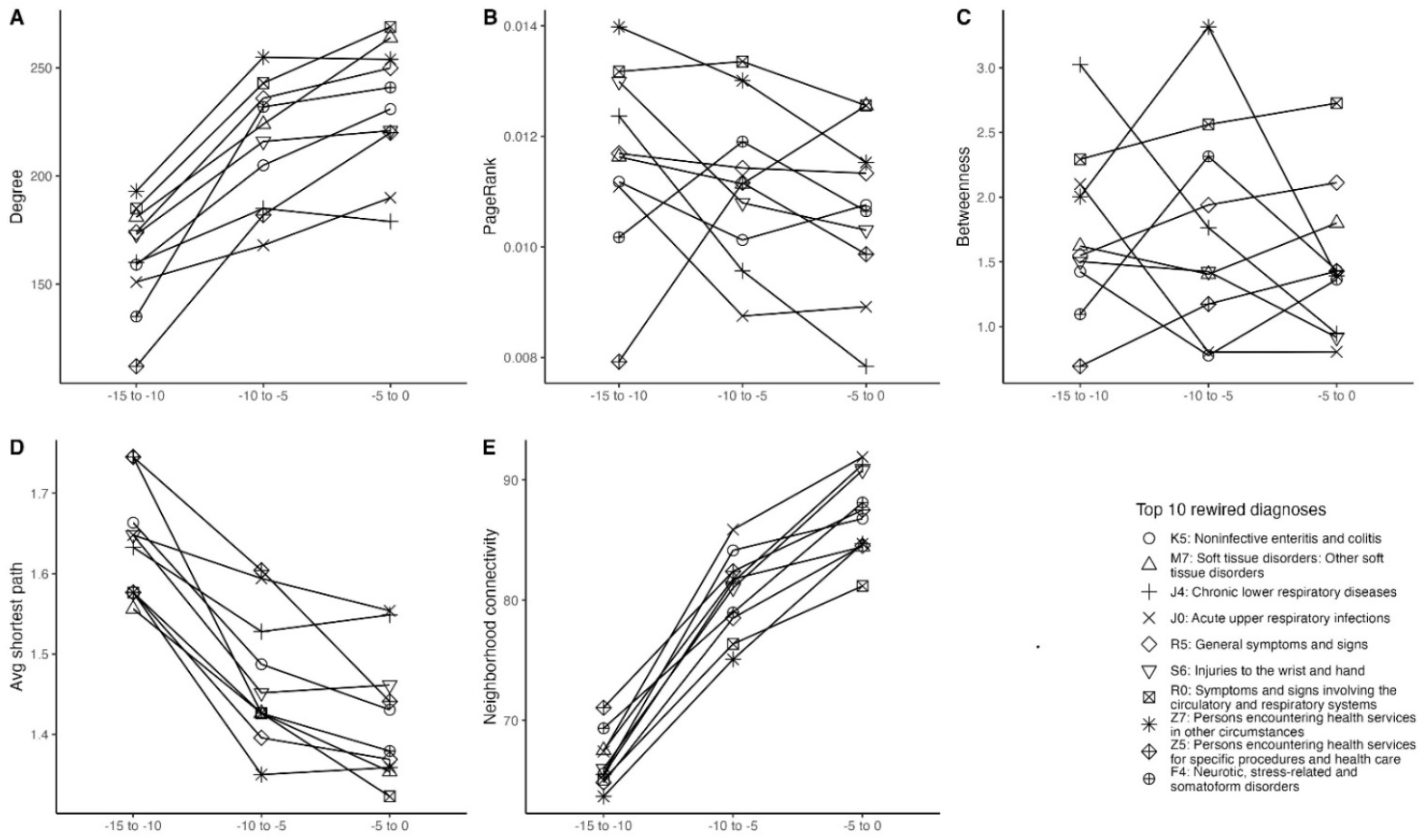
Comparison of centrality for the top 10 rewired diagnoses selected by DyNet score in Controls before the index date.

## 4. Discussion

We developed a network-based approach using outpatient ICD-10 trajectories to capture not only pre-onset diagnoses but their evolving interconnections. Compared with matched controls, the MS prodromal network was more compartmentalized, with fewer diagnoses and lower edge density (0.011 vs. 0.013), as well as lower mean degree, indicating fewer links per diagnosis. These patterns suggest that future MS patients follow more segmented diagnostic pathways, forming focused, modular clusters consistent with system-level disruption preceding onset.

Network clustering revealed clear differences between MS and controls. In MS, diagnoses formed multisystem communities spanning specialties, whereas control clusters were largely specialty-bounded (e.g., discrete ophthalmologic or dermatologic groups), reinforcing that the MS prodrome extends beyond neurology. Psychiatric comorbidities (depression, anxiety) were more frequent in the five years pre-onset and clustered tightly with other prodromal conditions (3, 22), consistent with reports of increased pre-diagnostic healthcare use, pain, psychiatric symptoms, and infections before MS diagnosis (3–9, 23–25). The prominence of mood and anxiety disorders, abdominal pain, musculoskeletal complaints, and injury-related encounters among central nodes mirrors prior findings, as does the endocrine–metabolic cluster including type 1 diabetes, hypothyroidism, and dyslipidaemia, aligning with evidence of polyautoimmunity and immune-mediated comorbidities in MS (7–9, 26). These results complement recent EHR-based studies of dynamic prodromal signatures (27), while extending them by focusing on diagnosis–diagnosis networks to characterize phenotypic clustering and rewiring.

Differences in healthcare use may partly explain denser pre-index MS networks, as more encounters yield more recorded diagnoses; however, encounter frequency alone cannot generate the structured co-occurrence patterns observed. The clustered MS networks therefore likely reflect both healthcare utilization and genuine prodromal features. In contrast to the largely isolated patterns in controls, MS prodromal networks formed interlinked constellations spanning mental health, metabolic, respiratory/allergic, and general symptom domains, consistent with systemic disruption before diagnosis.

An important contribution is the characterization of temporal reorganization in the MS prodrome. Longitudinal rewiring analysis captured shifts in diagnostic co-occurrence as onset approached, revealing changes in node centrality and inter-cluster bridging not detectable by frequency alone. Diagnoses trivial in isolation (e.g., general examination) gained late-prodromal relevance through repeated cross-cluster co-occurrence, indicating that structural position (hub vs. peripheral, bridge vs. isolated) represents an independent risk dimension. Consistent with signal aggregation, clustered weak indicators, such as fatigue, mood disturbance, viral infection, and allergic disease were more informative than any single symptom.

A system-level view of the MS prodrome emphasizes pattern recognition over isolated symptoms, suggesting that concurrent comorbidity clusters in younger or middle-aged adults should raise suspicion for MS before classical neurological signs. Rather than individual nonspecific complaints, the constellation itself may serve as a red flag, justifying closer monitoring, targeted testing, and earlier referral.

Our findings align with biomarker evidence implicating early inflammatory and infectious processes, including frequent viral infections, consistent with proposed roles of pathogens such as EBV (2). Although causality cannot be inferred, increased infectious-disease encounters in the prodrome may reflect exposures that trigger or accelerate autoimmunity. We also observed modest oncological and obstetric clustering, potentially reflecting known disease modifiers (e.g., pregnancy-related effects). Together, these patterns highlight the multisystem nature of the MS prodrome and warrant prospective validation.

From a systems-medicine perspective, integrating longitudinal EHRs with network analysis can reveal early-warning signals in complex diseases. Prior work in type 1 diabetes and cancer shows pre-onset network destabilization (28, 29); we demonstrate a comparable clinical-level phenomenon in MS, with diagnostic co-occurrence networks reorganizing as onset approaches. This suggests that EHRs could be monitored for prodromal network signatures, such as rapid clustering of autoimmune, psychiatric, and neurological diagnoses, to prompt earlier neurological evaluation. With prospective validation, network-based risk scores and visualizations could support primary-care decision-making.

This study has several limitations. Use of administrative NPR data entails potential ICD-10 misclassification, and comorbid diagnoses were not formally validated. Absence of primary care data likely under-ascertains milder conditions, biasing prodromal patterns toward more severe disease. Residual confounding remains possible due to unmeasured factors (e.g., lifestyle, health-seeking behaviour). Network findings may be context-specific to Swedish healthcare despite mitigation steps and require external replication. MS heterogeneity by sex, age, or phenotype was not examined. Finaly, Formal null-model testing was not performed, as suitable epidemiological null models for weighted disease networks are lacking; developing such frameworks remains an important methodological priority.

In summary, this case–control network analysis characterizes system-level diagnostic trajectories preceding MS. Mapping sequential transitions and rewiring reveal a prodromal architecture distinct from controls, with increasingly structured multisystem clusters spanning psychological, metabolic, immunologic, and general-symptom domains as onset approaches. These coordinated changes, largely absent in controls, suggest an evolving diagnostic complexity rather than isolated prodromal signals. While informative at the population level, this framework is hypothesis-generating and requires prospective validation and integration with clinical or biological markers before clinical use.

## 5. Statements & Declarations

### 5.1. Ethical considerations

This study involves human participants and was approved by the Regional Ethical Review Board at Karolinska Institutet (Dnr: 2017/1378-31), the Swedish Ethical Review Authority.

### 5.2. Consent to participate

Consent not required/was exempted, as approved by the Regional Ethical Review Board at Karolinska Institutet, the Swedish Ethical Review Authority. Specifically, the health information privacy regulations allow for the collection and analyses of healthcare and demographic data, without consent, for health-related research purposes. All data are stripped of identifiers before being released to preapproved researchers for analyses.

### 5.3. Consent for publication

Not applicable

### 5.6. Data availability

The data underlying this article were obtained from the Swedish National Patient Register (NPR). Restrictions apply to the availability of these data, which were used under license for this study. Data may be available from the Swedish National Board of Health and Welfare upon reasonable request and with appropriate approvals.

## References

1. Fagnani C, Neale MC, Nisticò L, Stazi MA, Ricigliano VA, Buscarinu MC, et al. Twin studies in multiple sclerosis: A meta-estimation of heritability and environmentality. Multiple sclerosis journal. 2015;21(11):1404–13.

2. Olsson T, Barcellos LF, Alfredsson L. Interactions between genetic, lifestyle and environmental risk factors for multiple sclerosis. Nature Reviews Neurology. 2017;13(1):25–36.

3. Wijnands JM, Kingwell E, Zhu F, Zhao Y, Högg T, Stadnyk K, et al. Health-care use before a first demyelinating event suggestive of a multiple sclerosis prodrome: a matched cohort study. The Lancet Neurology. 2017;16(6):445–51.

4. Thacker EL, Mirzaei F, Ascherio A. Infectious mononucleosis and risk for multiple sclerosis: a meta-analysis. Annals of neurology. 2006;59(3):499–503.

5. Montgomery S, Hiyoshi A, Burkill S, Alfredsson L, Bahmanyar S, Olsson T. Concussion in adolescence and risk of multiple sclerosis. Annals of neurology. 2017;82(4):554–61.

6. Povolo CA, Reid JN, Shariff SZ, Welk B, Morrow SA. Concussion in adolescence and the risk of multiple sclerosis: A retrospective cohort study. Multiple Sclerosis Journal. 2021;27(2):180–7.

7. Perga S, Martire S, Montarolo F, Giordani I, Spadaro M, Bono G, et al. The footprints of poly-autoimmunity: evidence for common biological factors involved in multiple sclerosis and Hashimoto’s thyroiditis. Frontiers in Immunology. 2018;9:311.

8. Islam MM, Poly TN, Yang H-C, Wu C-C, Li Y-C. Increase risk of multiple sclerosis in patients with psoriasis disease: an evidence of observational studies. Neuroepidemiology. 2019;52(3-4):152-60.

9. Kosmidou M, Katsanos AH, Katsanos KH, Kyritsis AP, Tsivgoulis G, Christodoulou D, et al. Multiple sclerosis and inflammatory bowel diseases: a systematic review and meta-analysis. Journal of neurology. 2017;264:254–9.

10. Gustafsson M, Nestor CE, Zhang H, Barabási A-L, Baranzini S, Brunak S, et al. Modules, networks and systems medicine for understanding disease and aiding diagnosis. Genome medicine. 2014;6(10):82.

11. Jensen PB, Jensen LJ, Brunak S. Mining electronic health records: towards better research applications and clinical care. Nature Reviews Genetics. 2012;13(6):395–405.

12. Ritchie MD, de Andrade M, Kuivaniemi H. The foundation of precision medicine: integration of electronic health records with genomics through basic, clinical, and translational research. Frontiers Media SA; 2015. p. 104.

13. Murley C, Friberg E, Hillert J, Alexanderson K, Yang F. Validation of multiple sclerosis diagnoses in the Swedish National Patient Register. European journal of epidemiology. 2019;34:1161–9.

14. Ludvigsson JF, Andersson E, Ekbom A, Feychting M, Kim J-L, Reuterwall C, et al. External review and validation of the Swedish national inpatient register. BMC public health. 2011;11(1):450.

15. Hillert J, Stawiarz L. The Swedish MS registry–clinical support tool and scientific resource. Acta Neurologica Scandinavica. 2015;132:11–9.

16. Alping P, Piehl F, Langer-Gould A, Frisell T. Validation of the Swedish multiple sclerosis register: further improving a resource for pharmacoepidemiologic evaluations. Epidemiology. 2019;30(2):230–3.

17. Tremlett H, Zhu F, Everett K, Asaf A, Manouchehrinia A, Li P, et al. Healthcare use is elevated two decades before a first demyelinating event and differs by age and sex. Annals of Clinical and Translational Neurology. 2025;12(2):415–32.

18. Manouchehrinia A, Zhu F, Hillert J, McKay K, Zhao Y, Marrie RA, et al. Prodromal phase of multiple sclerosis: evidence from sickness absence patterns before disease onset–a matched cohort study. Journal of Neurology, Neurosurgery & Psychiatry. 2025.

19. Morris JH, Apeltsin L, Newman AM, Baumbach J, Wittkop T, Su G, et al. clusterMaker: a multi-algorithm clustering plugin for Cytoscape. BMC bioinformatics. 2011;12(1):436.

20. Goenawan IH, Bryan K, Lynn DJ. DyNet: visualization and analysis of dynamic molecular interaction networks. Bioinformatics. 2016;32(17):2713–5.

21. Franz M, Lopes CT, Huck G, Dong Y, Sumer O, Bader GD. Cytoscape. js: a graph theory library for visualisation and analysis. Bioinformatics. 2016;32(2):309–11.

22. Chertcoff AS, Yusuf FL, Zhu F, Evans C, Fisk JD, Zhao Y, et al. Psychiatric comorbidity during the prodromal period in patients with multiple sclerosis. Neurology. 2023;101(20):e2026–e34.

23. Makhani N, Tremlett H. The multiple sclerosis prodrome. Nature Reviews Neurology. 2021;17(8):515–21.

24. Disanto G, Zecca C, MacLachlan S, Sacco R, Handunnetthi L, Meier UC, et al. Prodromal symptoms of multiple sclerosis in primary care. Annals of neurology. 2018;83(6):1162–73.

25. Tremlett H, Marrie RA. The multiple sclerosis prodrome: Emerging evidence, challenges, and opportunities. Multiple Sclerosis Journal. 2021;27(1):6–12.

26. Deretzi G, Kountouras J, Polyzos S, Koutlas E, Pelidou SH, Xeromerisiou G, et al. Polyautoimmunity in a Greek cohort of multiple sclerosis. Acta Neurologica Scandinavica. 2015;131(4):225–30.

27. Nelson CA, Bove R, Butte AJ, Baranzini SE. Embedding electronic health records onto a knowledge network recognizes prodromal features of multiple sclerosis and predicts diagnosis. Journal of the American Medical Informatics Association. 2022;29(3):424–34.

28. Han Y, Akhtar J, Liu G, Li C, Wang G. Early warning and diagnosis of liver cancer based on dynamic network biomarker and deep learning. Computational and Structural Biotechnology Journal. 2023;21:3478–89.

29. Liu X, Liu R, Zhao X-M, Chen L. Detecting early-warning signals of type 1 diabetes and its leading biomolecular networks by dynamical network biomarkers. BMC medical genomics. 2013;6(Suppl 2):S8.

