## Supplementary Materials for "A susceptibility network analysis of disease trajectories leading to multiple sclerosis: a nationwide cohort study"

Table S 1: Comparison of topological network measures between MS and non-MS

|  | Control (N=30008) | MS (N=29560) | Total (N=59568) | p value |
| --- | --- | --- | --- | --- |
| **Degree** | | | | <0.001 |
| Median (Q1, Q3) | 0.021 (0.006, 0.065) | 0.022 (0.007, 0.070) | 0.022 (0.006, 0.068) |  |
| Min | 0.001 | 0.001 | 0.001 |  |
| Max | 0.818 | 0.815 | 0.818 |  |
| **Closeness** | | | | <0.001 |
| Median (Q1, Q3) | 75717.773 (67361.534, 82116.950) | 78084.458 (69362.148, 84531.683) | 76898.209 (68472.261, 83316.919) |  |
| Min | 16680.976 | 42967.208 | 16680.976 |  |
| Max | 98364.541 | 95762.104 | 98364.541 |  |
| **Betweenness** | | | | 0.950 |
| Median (Q1, Q3) | 0.000 (0.000, 0.001) | 0.000 (0.000, 0.001) | 0.000 (0.000, 0.001) |  |
| Min | 0.000 | 0.000 | 0.000 |  |
| Max | 0.039 | 0.032 | 0.039 |  |
| **Eigenvalue** | | | | <0.001 |
| Median (Q1, Q3) | 0.001 (0.000, 0.004) | 0.002 (0.000, 0.007) | 0.001 (0.000, 0.005) |  |
| Min | 0.000 | 0.000 | 0.000 |  |
| Max | 1.000 | 1.000 | 1.000 |  |
| **pagerank** |  | | | 0.326 |
| Median (Q1, Q3) | 0.000 (0.000, 0.001) | 0.000 (0.000, 0.001) | 0.000 (0.000, 0.001) |  |
| Min | 0.000 | 0.000 | 0.000 |  |
| Max | 0.019 | 0.019 | 0.019 |  |
| **transitivity_local** | | | | <0.001 |
| Median (Q1, Q3) | 0.393 (0.320, 0.500) | 0.427 (0.335, 0.519) | 0.409 (0.331, 0.503) |  |
| Min | 0.000 | 0.000 | 0.000 |  |
| Max | 1.000 | 1.000 | 1.000 |  |
| **transitivity_global** | | | | <0.001 |
| Median (Q1, Q3) | 0.288 (0.287, 0.289) | 0.299 (0.299, 0.299) | 0.291 (0.288, 0.299) |  |
| Min | 0.284 | 0.299 | 0.284 |  |
| Max | 0.291 | 0.299 | 0.299 |  |

Table S 2: Centrality measures of the top 30 nodes in each networks

| **MS patients** | | | | | | | | | |
| --- | --- | --- | --- | --- | --- | --- | --- | --- | --- |
| **DC** | | **CC** | | **BC** | | **EC** | | **PR** | |
| ICD | Metric value | ICD | Metric value | ICD | Metric value | ICD | Metric value | ICD | Metric value |
| R10 | 0.815166 | J45 | 95762.1 | R10 | 0.031775 | R10 | 1 | Z03 | 0.018516 |
| Z03 | 0.752877 | Z03 | 95727.73 | Z03 | 0.029314 | Z03 | 0.782013 | R10 | 0.018467 |
| Z01 | 0.729858 | R07 | 95727.73 | Z01 | 0.023942 | F41 | 0.474724 | Z09 | 0.014955 |
| Z09 | 0.680433 | N39 | 95385.36 | Z09 | 0.022848 | F32 | 0.405745 | Z01 | 0.014812 |
| M79 | 0.560596 | M79 | 95283.12 | S06 | 0.021867 | Z01 | 0.40397 | O80 | 0.010594 |
| M54 | 0.505078 | S06 | 95113.22 | M79 | 0.017747 | E10 | 0.395681 | E10 | 0.010112 |
| O80 | 0.498307 | M54 | 94842.62 | J45 | 0.016657 | Z09 | 0.271468 | F41 | 0.009951 |
| Z71 | 0.484089 | Z01 | 94775.22 | F32 | 0.016108 | R20 | 0.265843 | R20 | 0.009268 |
| R07 | 0.471225 | R42 | 94607.12 | Z71 | 0.015974 | O80 | 0.22112 | M79 | 0.009215 |
| R20 | 0.462424 | N92 | 94607.12 | N39 | 0.015563 | M79 | 0.220638 | F32 | 0.008772 |
| J45 | 0.459716 | R51 | 94607.12 | M54 | 0.015476 | Z34 | 0.180973 | M54 | 0.007677 |
| N39 | 0.454976 | Z09 | 94540.04 | R07 | 0.015409 | F10 | 0.168728 | J45 | 0.006765 |
| F32 | 0.43128 | R10 | 94473.06 | O80 | 0.014511 | M54 | 0.167488 | Z30 | 0.006107 |
| F41 | 0.425863 | R20 | 94473.06 | I10 | 0.014105 | N97 | 0.163929 | Z51 | 0.006058 |
| Z30 | 0.417062 | Z30 | 94239.38 | A41 | 0.013891 | F90 | 0.163182 | I10 | 0.005789 |
| N92 | 0.412322 | K35 | 94239.38 | R20 | 0.01385 | F43 | 0.161011 | Z71 | 0.005706 |
| I10 | 0.394042 | F32 | 94073.18 | N92 | 0.013313 | F31 | 0.151094 | N92 | 0.005641 |
| E10 | 0.391334 | Z71 | 94040.01 | E10 | 0.012964 | Z30 | 0.147959 | N97 | 0.005487 |
| S06 | 0.383886 | S52 | 93841.47 | F41 | 0.01256 | Z71 | 0.138798 | F10 | 0.005421 |
| R42 | 0.374408 | D22 | 93709.58 | A09 | 0.012364 | F60 | 0.135141 | Z34 | 0.005289 |
| D22 | 0.354773 | S61 | 93709.58 | O03 | 0.012151 | N92 | 0.129077 | N39 | 0.005223 |
| R51 | 0.347326 | O80 | 93610.9 | Z30 | 0.011861 | R07 | 0.128521 | F90 | 0.005197 |
| E66 | 0.327691 | I10 | 93610.9 | J06 | 0.010819 | F33 | 0.128065 | R07 | 0.005097 |
| F43 | 0.323629 | A41 | 93578.06 | R42 | 0.010734 | N83 | 0.121355 | F31 | 0.005053 |
| G40 | 0.317536 | M25 | 93250.86 | K80 | 0.010409 | O26 | 0.117703 | G40 | 0.004935 |
| K80 | 0.31415 | G56 | 93218.27 | K35 | 0.010307 | J45 | 0.115834 | F43 | 0.004854 |
| Z64 | 0.313473 | K80 | 93185.69 | F10 | 0.010296 | K80 | 0.106825 | R42 | 0.004598 |
| M25 | 0.311442 | Z04 | 93153.15 | N20 | 0.009819 | N39 | 0.10552 | O26 | 0.004582 |
| F10 | 0.309411 | J06 | 92958.33 | S82 | 0.009608 | S82 | 0.098722 | H20 | 0.004408 |
| R52 | 0.308734 | J30 | 92925.94 | F43 | 0.009303 | R42 | 0.098635 | H53 | 0.004401 |
| Control | | | | | | | | | |
| **DC** | | **CC** | | **BC** | | **EC** | | **PR** | |
| ICD | Metric | ICD | Metric | ICD | Metric | ICD | Metric | ICD | Metric |
| R10 | 0.816911 | Z01 | 101408.6 | R10 | 0.041111 | R10 | 1 | R10 | 0.019034 |
| Z09 | 0.695941 | Z03 | 100902.5 | Z01 | 0.03254 | F10 | 1 | Z09 | 0.01718 |
| Z01 | 0.678146 | R07 | 99623.86 | Z03 | 0.031983 | C50 | 1 | O80 | 0.013685 |
| Z03 | 0.6 | N39 | 99274.42 | Z09 | 0.03088 | E10 | 1 | Z01 | 0.011864 |
| M79 | 0.52969 | R10 | 99031.27 | R07 | 0.023893 | Z51 | 0.987617 | E10 | 0.01166 |
| O80 | 0.518912 | Z71 | 98754.84 | S06 | 0.023634 | F41 | 0.933683 | F41 | 0.010282 |
| R07 | 0.493333 | S06 | 98754.84 | N39 | 0.022453 | N97 | 0.831843 | F10 | 0.010026 |
| Z71 | 0.468043 | M54 | 98582.86 | E10 | 0.02118 | F32 | 0.797876 | Z51 | 0.009328 |
| I10 | 0.455629 | M79 | 98343.08 | A41 | 0.021035 | F60 | 0.424552 | M79 | 0.008644 |
| M54 | 0.454 | J45 | 98104.46 | I10 | 0.020825 | O80 | 0.357055 | F32 | 0.008604 |
| J45 | 0.454 | R06 | 98070.47 | M79 | 0.020496 | Z09 | 0.310894 | I10 | 0.008577 |
| E10 | 0.449767 | K80 | 97900.86 | J45 | 0.019765 | Z31 | 0.302972 | Z03 | 0.008135 |
| N39 | 0.444 | O80 | 97563.39 | M54 | 0.019277 | F43 | 0.301344 | J45 | 0.007801 |
| F41 | 0.442645 | I10 | 97228.23 | O80 | 0.018267 | Z01 | 0.2835 | Z30 | 0.007125 |
| Z30 | 0.430883 | A41 | 97161.48 | F32 | 0.018158 | F33 | 0.24507 | C50 | 0.006963 |
| F32 | 0.421727 | S52 | 97061.52 | F41 | 0.017393 | F50 | 0.229707 | M54 | 0.006589 |
| S06 | 0.398111 | Z09 | 96862.21 | A09 | 0.016625 | H36 | 0.224265 | F60 | 0.006495 |
| N92 | 0.390812 | F32 | 96729.79 | Z71 | 0.016536 | Z34 | 0.203808 | N97 | 0.006459 |
| D22 | 0.373085 | K35 | 96696.75 | T81 | 0.01625 | J45 | 0.196787 | G40 | 0.005925 |
| F10 | 0.368421 | Z30 | 96663.72 | K35 | 0.015757 | F31 | 0.196782 | Z08 | 0.005729 |
| Z64 | 0.334893 | S83 | 96630.72 | F10 | 0.0156 | J30 | 0.196611 | R07 | 0.005704 |
| F43 | 0.332881 | N83 | 96630.72 | Z30 | 0.015266 | M79 | 0.193112 | F43 | 0.005606 |
| E66 | 0.331126 | D22 | 96630.72 | J06 | 0.014893 | Z30 | 0.190005 | Z34 | 0.005599 |
| K80 | 0.328676 | K40 | 96564.78 | S82 | 0.013374 | Z03 | 0.188507 | M05 | 0.005532 |
| O03 | 0.325566 | J06 | 96498.94 | D22 | 0.013196 | Z08 | 0.180294 | N92 | 0.005484 |
| T81 | 0.322193 | A09 | 96466.05 | R06 | 0.012948 | M05 | 0.167025 | Z71 | 0.00548 |
| Z51 | 0.313907 | S62 | 96466.05 | O03 | 0.012904 | C77 | 0.161569 | N39 | 0.005422 |
| K35 | 0.307797 | R52 | 96466.05 | K80 | 0.012791 | O26 | 0.159042 | M32 | 0.005195 |
| S52 | 0.306 | Z64 | 96466.05 | S83 | 0.012492 | M54 | 0.151912 | E11 | 0.005191 |
| A09 | 0.305085 | K59 | 96334.71 | N92 | 0.012454 | I10 | 0.146895 | L40 | 0.005065 |

Table S 3: Clusters heterogeneity summary

| group | avg_shannon | avg_n_groups |
| --- | --- | --- |
| Control | 1.641403252 | 7.7 |
| MS | 2.024083545 | 12.6 |

Table S 4: Cluster heterogeneity per MS and Control clusters

| group | cluster | dominant_group | dominant_count | total_icds | shannon_H | n_groups |
| --- | --- | --- | --- | --- | --- | --- |
| MS | 1 | R | 22 | 142 | 2.762142683 | 20 |
| MS | 2 | S | 34 | 106 | 2.406143344 | 18 |
| MS | 3 | K | 21 | 89 | 2.476299071 | 18 |
| MS | 4 | E | 9 | 45 | 2.575734712 | 16 |
| MS | 5 | C | 17 | 42 | 1.912278338 | 11 |
| MS | 6 | F | 17 | 35 | 1.822740173 | 12 |
| MS | 7 | O | 21 | 30 | 0.955582211 | 5 |
| MS | 8 | F | 8 | 25 | 2.178884033 | 12 |
| MS | 9 | F | 10 | 20 | 1.541018448 | 7 |
| MS | 10 | J | 8 | 17 | 1.610012437 | 7 |
| Control | 1 | S | 70 | 808 | 2.965424427 | 23 |
| Control | 2 | Q | 6 | 29 | 2.175218847 | 11 |
| Control | 3 | L | 6 | 14 | 1.673118363 | 7 |
| Control | 4 | H | 6 | 13 | 1.311431337 | 5 |
| Control | 5 | N | 9 | 12 | 0.836988217 | 4 |
| Control | 6 | G | 2 | 11 | 2.019814992 | 8 |
| Control | 7 | H | 4 | 10 | 1.279854226 | 4 |
| Control | 8 | L | 6 | 10 | 1.088899975 | 4 |
| Control | 9 | K | 3 | 9 | 1.676987774 | 6 |
| Control | 10 | B | 4 | 8 | 1.386294361 | 5 |

Table S 5: Control clusters taglines

| cluster | icd_group | icd_group_name | top_conditions | tagline |
| --- | --- | --- | --- | --- |
| 1 | S | Injury, poisoning and certain other consequences of external causes | Dislocation, sprain and strain of joints and ligaments of knee; Fracture of forearm; Fracture at wrist and hand level; Fracture of lower leg, including ankle; Open wound of wrist and hand | Cluster 1 (S: Injury, poisoning and certain other consequences of external causes): Dislocation, sprain and strain of joints and ligaments of knee; Fracture of forearm; Fracture at wrist and hand level; Fracture of lower leg, including ankle; Open wound of wrist and hand |
| 2 | Q | Congenital malformations, deformations and chromosomal abnormalities | Phakomatoses, not elsewhere classified; Congenital obstructive defects of renal pelvis and congenital malformations of ureter; Other congenital malformations of circulatory system; Monosomies and deletions from the autosomes, not elsewhere classified; Other chromosome abnormalities, not elsewhere classified | Cluster 2 (Q: Congenital malformations, deformations and chromosomal abnormalities): Phakomatoses, not elsewhere classified; Congenital obstructive defects of renal pelvis and congenital malformations of ureter; Other congenital malformations of circulatory system; Monosomies and deletions from the autosomes, not elsewhere classified; Other chromosome abnormalities, not elsewhere classified |
| 3 | L | Diseases of the skin and subcutaneous tissue | Acne; Seborrhoeic keratosis; Other disorders of pigmentation; Seborrhoeic dermatitis; Other acute skin changes due to ultraviolet radiation | Cluster 3 (L: Diseases of the skin and subcutaneous tissue): Acne; Seborrhoeic keratosis; Other disorders of pigmentation; Seborrhoeic dermatitis; Other acute skin changes due to ultraviolet radiation |
| 4 | H | Diseases of the eye and adnexa / ear and mastoid process | Disorders of refraction and accommodation; Other strabismus; Visual disturbances; Visual impairment including blindness (binocular or monocular); Nystagmus and other irregular eye movements | Cluster 4 (H: Diseases of the eye and adnexa / ear and mastoid process): Disorders of refraction and accommodation; Other strabismus; Visual disturbances; Visual impairment including blindness (binocular or monocular); Nystagmus and other irregular eye movements |
| 5 | N | Diseases of the genitourinary system | Chronic kidney disease; Obstructive and reflux uropathy; Chronic tubulo-interstitial nephritis; Recurrent and persistent haematuria; Hereditary nephropathy, not elsewhere classified | Cluster 5 (N: Diseases of the genitourinary system): Chronic kidney disease; Obstructive and reflux uropathy; Chronic tubulo-interstitial nephritis; Recurrent and persistent haematuria; Hereditary nephropathy, not elsewhere classified |
| 6 | G | Diseases of the nervous system | Inflammatory polyneuropathy; Vascular syndromes of brain in cerebrovascular diseases (I60-I67+) | Cluster 6 (G: Diseases of the nervous system): Inflammatory polyneuropathy; Vascular syndromes of brain in cerebrovascular diseases (I60-I67+) |
| 7 | H | Diseases of the eye and adnexa / ear and mastoid process | Keratitis; Disorders of lacrimal system; Other inflammation of eyelid; Corneal scars and opacities | Cluster 7 (H: Diseases of the eye and adnexa / ear and mastoid process): Keratitis; Disorders of lacrimal system; Other inflammation of eyelid; Corneal scars and opacities |
| 8 | L | Diseases of the skin and subcutaneous tissue | Other dermatitis; Allergic contact dermatitis; Lichen simplex chronicus and prurigo; Unspecified contact dermatitis; Impetigo | Cluster 8 (L: Diseases of the skin and subcutaneous tissue): Other dermatitis; Allergic contact dermatitis; Lichen simplex chronicus and prurigo; Unspecified contact dermatitis; Impetigo |
| 9 | K | Diseases of the digestive system | Ulcerative colitis; Other noninfective gastroenteritis and colitis; Other diseases of biliary tract | Cluster 9 (K: Diseases of the digestive system): Ulcerative colitis; Other noninfective gastroenteritis and colitis; Other diseases of biliary tract |
| 10 | B | Certain infectious and parasitic diseases | Chronic viral hepatitis; Unspecified human immunodeficiency virus [HIV] disease; Acute hepatitis B; Unspecified viral hepatitis | Cluster 10 (B: Certain infectious and parasitic diseases): Chronic viral hepatitis; Unspecified human immunodeficiency virus [HIV] disease; Acute hepatitis B; Unspecified viral hepatitis |

Table S 6: MS clusters taglines

| cluster | icd_group | icd_group_name | top_conditions | tagline |
| --- | --- | --- | --- | --- |
| 1 | R | Symptoms, signs and abnormal clinical and lab findings | Dizziness and giddiness; Headache; Pain in throat and chest; Abnormalities of breathing; Abnormalities of heart beat | Cluster 1 (R: Symptoms, signs and abnormal clinical and lab findings): Dizziness and giddiness; Headache; Pain in throat and chest; Abnormalities of breathing; Abnormalities of heart beat |
| 2 | S | Injury, poisoning and certain other consequences of external causes | Fracture of lower leg, including ankle; Fracture of forearm; Fracture of foot, except ankle; Dislocation, sprain and strain of joints and ligaments at wrist and hand level; Fracture of shoulder and upper arm | Cluster 2 (S: Injury, poisoning and certain other consequences of external causes): Fracture of lower leg, including ankle; Fracture of forearm; Fracture of foot, except ankle; Dislocation, sprain and strain of joints and ligaments at wrist and hand level; Fracture of shoulder and upper arm |
| 3 | K | Diseases of the digestive system | Cholelithiasis; Gastro-oesophageal reflux disease; Irritable bowel syndrome; Diaphragmatic hernia; Other functional intestinal disorders | Cluster 3 (K: Diseases of the digestive system): Cholelithiasis; Gastro-oesophageal reflux disease; Irritable bowel syndrome; Diaphragmatic hernia; Other functional intestinal disorders |
| 4 | E | Endocrine, nutritional and metabolic diseases | Type 1 diabetes mellitus; Type 2 diabetes mellitus; Other hypothyroidism; Disorders of lipoprotein metabolism and other lipidaemias; Unspecified diabetes mellitus | Cluster 4 (E: Endocrine, nutritional and metabolic diseases): Type 1 diabetes mellitus; Type 2 diabetes mellitus; Other hypothyroidism; Disorders of lipoprotein metabolism and other lipidaemias; Unspecified diabetes mellitus |
| 5 | C | Neoplasms | Malignant neoplasm of breast; Secondary and unspecified malignant neoplasm of lymph nodes; Malignant neoplasm of corpus uteri; Malignant neoplasm of testis; Malignant neoplasm of base of tongue | Cluster 5 (C: Neoplasms): Malignant neoplasm of breast; Secondary and unspecified malignant neoplasm of lymph nodes; Malignant neoplasm of corpus uteri; Malignant neoplasm of testis; Malignant neoplasm of base of tongue |
| 6 | F | Mental and behavioural disorders | Other anxiety disorders; Specific personality disorders; Mental and behavioural disorders due to use of alcohol; Pervasive developmental disorders; Phobic anxiety disorders | Cluster 6 (F: Mental and behavioural disorders): Other anxiety disorders; Specific personality disorders; Mental and behavioural disorders due to use of alcohol; Pervasive developmental disorders; Phobic anxiety disorders |
| 7 | O | Pregnancy, childbirth and the puerperium | Maternal care for other conditions predominantly related to pregnancy; False labour; Haemorrhage in early pregnancy; Other maternal diseases classifiable elsewhere but complicating pregnancy, childbirth and the puerperium; Maternal care for known or suspected malpresentation of fetus | Cluster 7 (O: Pregnancy, childbirth and the puerperium): Maternal care for other conditions predominantly related to pregnancy; False labour; Haemorrhage in early pregnancy; Other maternal diseases classifiable elsewhere but complicating pregnancy, childbirth and the puerperium; Maternal care for known or suspected malpresentation of fetus |
| 8 | F | Mental and behavioural disorders | Depressive episode; Somatoform disorders; Nonorganic sleep disorders; Mixed and other personality disorders; Other mood [affective] disorders | Cluster 8 (F: Mental and behavioural disorders): Depressive episode; Somatoform disorders; Nonorganic sleep disorders; Mixed and other personality disorders; Other mood [affective] disorders |
| 9 | F | Mental and behavioural disorders | Hyperkinetic disorders; Mental and behavioural disorders due to use of other stimulants, including caffeine; Other behavioural and emotional disorders with onset usually occurring in childhood and adolescence; Mental and behavioural disorders due to use of cannabinoids; Dissociative [conversion] disorders | Cluster 9 (F:Mental and behavioural disorders): Hyperkinetic disorders; Mental and behavioural disorders due to use of other stimulants, including caffeine; Other behavioural and emotional disorders with onset usually occurring in childhood and adolescence; Mental and behavioural disorders due to use of cannabinoids; Dissociative [conversion] disorders |
| 10 | J | Diseases of the respiratory system | Asthma; Vasomotor and allergic rhinitis; Other respiratory disorders; Other chronic obstructive pulmonary disease; Other diseases of upper respiratory tract | Cluster 10 (J: Diseases of the respiratory system): Asthma; Vasomotor and allergic rhinitis; Other respiratory disorders; Other chronic obstructive pulmonary disease; Other diseases of upper respiratory tract |

Table S 7: Top 5 most strength nodes in each Control clusters

| cluster | icd_group | icd_group_name | icd_code | label | strength | degree |
| --- | --- | --- | --- | --- | --- | --- |
| 1 | S | Injury, poisoning and certain other consequences of external causes | S83 | Dislocation, sprain and strain of joints and ligaments of knee | 0.006278 | 70 |
| 1 | S | Injury, poisoning and certain other consequences of external causes | S52 | Fracture of forearm | 0.006272 | 69 |
| 1 | S | Injury, poisoning and certain other consequences of external causes | S62 | Fracture at wrist and hand level | 0.006176 | 76 |
| 1 | S | Injury, poisoning and certain other consequences of external causes | S82 | Fracture of lower leg, including ankle | 0.004549 | 62 |
| 1 | S | Injury, poisoning and certain other consequences of external causes | S61 | Open wound of wrist and hand | 0.002513 | 83 |
| 2 | Q | Congenital malformations, deformations and chromosomal abnormalities | Q85 | Phakomatoses, not elsewhere classified | 0.000242 | 2 |
| 2 | Q | Congenital malformations, deformations and chromosomal abnormalities | Q62 | Congenital obstructive defects of renal pelvis and congenital malformations of ureter | 3.12E-05 | 2 |
| 2 | Q | Congenital malformations, deformations and chromosomal abnormalities | Q28 | Other congenital malformations of circulatory system | 2.34E-05 | 2 |
| 2 | Q | Congenital malformations, deformations and chromosomal abnormalities | Q93 | Monosomies and deletions from the autosomes, not elsewhere classified | 2.14E-05 | 3 |
| 2 | Q | Congenital malformations, deformations and chromosomal abnormalities | Q99 | Other chromosome abnormalities, not elsewhere classified | 9.74E-06 | 3 |
| 3 | L | Diseases of the skin and subcutaneous tissue | L70 | Acne | 0.003026 | 10 |
| 3 | L | Diseases of the skin and subcutaneous tissue | L82 | Seborrhoeic keratosis | 0.000314 | 11 |
| 3 | L | Diseases of the skin and subcutaneous tissue | L81 | Other disorders of pigmentation | 0.000247 | 7 |
| 3 | L | Diseases of the skin and subcutaneous tissue | L21 | Seborrhoeic dermatitis | 0.000158 | 8 |
| 3 | L | Diseases of the skin and subcutaneous tissue | L56 | Other acute skin changes due to ultraviolet radiation | 4.09E-05 | 5 |
| 4 | H | Diseases of the eye and adnexa / ear and mastoid process | H52 | Disorders of refraction and accommodation | 0.002697 | 12 |
| 4 | H | Diseases of the eye and adnexa / ear and mastoid process | H50 | Other strabismus | 0.001752 | 11 |
| 4 | H | Diseases of the eye and adnexa / ear and mastoid process | H53 | Visual disturbances | 0.000711 | 11 |
| 4 | H | Diseases of the eye and adnexa / ear and mastoid process | H54 | Visual impairment including blindness (binocular or monocular) | 0.000127 | 9 |
| 4 | H | Diseases of the eye and adnexa / ear and mastoid process | H55 | Nystagmus and other irregular eye movements | 0.000111 | 10 |
| 5 | N | Diseases of the genitourinary system | N18 | Chronic kidney disease | 0.001621 | 14 |
| 5 | N | Diseases of the genitourinary system | N13 | Obstructive and reflux uropathy | 0.000452 | 8 |
| 5 | N | Diseases of the genitourinary system | N11 | Chronic tubulo-interstitial nephritis | 0.000166 | 6 |
| 5 | N | Diseases of the genitourinary system | N02 | Recurrent and persistent haematuria | 8.38E-05 | 4 |
| 5 | N | Diseases of the genitourinary system | N07 | Hereditary nephropathy, not elsewhere classified | 7.99E-05 | 6 |
| 6 | G | Diseases of the nervous system | G61 | Inflammatory polyneuropathy | 0.000148 | 2 |
| 6 | G | Diseases of the nervous system | G46 | Vascular syndromes of brain in cerebrovascular diseases (I60-I67+) | 0 | 0 |
| 7 | H | Diseases of the eye and adnexa / ear and mastoid process | H16 | Keratitis | 0.00324 | 8 |
| 7 | H | Diseases of the eye and adnexa / ear and mastoid process | H04 | Disorders of lacrimal system | 0.000922 | 8 |
| 7 | H | Diseases of the eye and adnexa / ear and mastoid process | H01 | Other inflammation of eyelid | 0.000284 | 7 |
| 7 | H | Diseases of the eye and adnexa / ear and mastoid process | H17 | Corneal scars and opacities | 0.000129 | 7 |
| 8 | L | Diseases of the skin and subcutaneous tissue | L30 | Other dermatitis | 0.002426 | 12 |
| 8 | L | Diseases of the skin and subcutaneous tissue | L23 | Allergic contact dermatitis | 0.000992 | 10 |
| 8 | L | Diseases of the skin and subcutaneous tissue | L28 | Lichen simplex chronicus and prurigo | 0.000173 | 7 |
| 8 | L | Diseases of the skin and subcutaneous tissue | L25 | Unspecified contact dermatitis | 0.000113 | 8 |
| 8 | L | Diseases of the skin and subcutaneous tissue | L01 | Impetigo | 0.000107 | 5 |
| 9 | K | Diseases of the digestive system | K51 | Ulcerative colitis | 0.005929 | 6 |
| 9 | K | Diseases of the digestive system | K52 | Other noninfective gastroenteritis and colitis | 0.000871 | 6 |
| 9 | K | Diseases of the digestive system | K83 | Other diseases of biliary tract | 0.000421 | 6 |
| 10 | B | Certain infectious and parasitic diseases | B18 | Chronic viral hepatitis | 0.001629 | 7 |
| 10 | B | Certain infectious and parasitic diseases | B24 | Unspecified human immunodeficiency virus [HIV] disease | 6.23E-05 | 4 |
| 10 | B | Certain infectious and parasitic diseases | B16 | Acute hepatitis B | 4.87E-05 | 5 |
| 10 | B | Certain infectious and parasitic diseases | B19 | Unspecified viral hepatitis | 1.36E-05 | 4 |

Table S 8: Top 5 most strength nodes in each MS clusters

| cluster | icd_group | icd_group_name | icd_code | label | strength | degree |
| --- | --- | --- | --- | --- | --- | --- |
| 1 | R | Symptoms, signs and abnormal clinical and lab findings | R42 | Dizziness and giddiness | 0.2616193955 | 21 |
| 1 | R | Symptoms, signs and abnormal clinical and lab findings | R51 | Headache | 0.2349730375 | 17 |
| 1 | R | Symptoms, signs and abnormal clinical and lab findings | R07 | Pain in throat and chest | 0.1477665445 | 19 |
| 1 | R | Symptoms, signs and abnormal clinical and lab findings | R06 | Abnormalities of breathing | 0.0855915485 | 17 |
| 1 | R | Symptoms, signs and abnormal clinical and lab findings | R00 | Abnormalities of heart beat | 0.062982453 | 13 |
| 2 | S | Injury, poisoning and certain other consequences of external causes | S82 | Fracture of lower leg, including ankle | 0.4893253275 | 32 |
| 2 | S | Injury, poisoning and certain other consequences of external causes | S52 | Fracture of forearm | 0.4618713325 | 33 |
| 2 | S | Injury, poisoning and certain other consequences of external causes | S92 | Fracture of foot, except ankle | 0.197022059 | 27 |
| 2 | S | Injury, poisoning and certain other consequences of external causes | S63 | Dislocation, sprain and strain of joints and ligaments at wrist and hand level | 0.1719905635 | 26 |
| 2 | S | Injury, poisoning and certain other consequences of external causes | S42 | Fracture of shoulder and upper arm | 0.171183093 | 30 |
| 3 | K | Diseases of the digestive system | K80 | Cholelithiasis | 0.1429217475 | 17 |
| 3 | K | Diseases of the digestive system | K21 | Gastro-oesophageal reflux disease | 0.1308097215 | 21 |
| 3 | K | Diseases of the digestive system | K58 | Irritable bowel syndrome | 0.105778236 | 14 |
| 3 | K | Diseases of the digestive system | K44 | Diaphragmatic hernia | 0.0985110305 | 22 |
| 3 | K | Diseases of the digestive system | K59 | Other functional intestinal disorders | 0.0928587665 | 16 |
| 4 | E | Endocrine, nutritional and metabolic diseases | E10 | Type 1 diabetes mellitus | ######## | 16 |
| 4 | E | Endocrine, nutritional and metabolic diseases | E11 | Type 2 diabetes mellitus | 0.21397887 | 9 |
| 4 | E | Endocrine, nutritional and metabolic diseases | E03 | Other hypothyroidism | 0.1154678345 | 11 |
| 4 | E | Endocrine, nutritional and metabolic diseases | E78 | Disorders of lipoprotein metabolism and other lipidaemias | 0.081554214 | 8 |
| 4 | E | Endocrine, nutritional and metabolic diseases | E14 | Unspecified diabetes mellitus | 0.07670941 | 6 |
| 5 | C | Neoplasms | C50 | Malignant neoplasm of breast | 0.505474716 | 8 |
| 5 | C | Neoplasms | C77 | Secondary and unspecified malignant neoplasm of lymph nodes | 0.055715254 | 9 |
| 5 | C | Neoplasms | C54 | Malignant neoplasm of corpus uteri | 0.05167792 | 2 |
| 5 | C | Neoplasms | C62 | Malignant neoplasm of testis | 0.0411808325 | 6 |
| 5 | C | Neoplasms | C01 | Malignant neoplasm of base of tongue | 0.04037338 | 2 |
| 6 | F | Mental and behavioural disorders | F41 | Other anxiety disorders | ######## | 27 |
| 6 | F | Mental and behavioural disorders | F60 | Specific personality disorders | 0.5805690105 | 15 |
| 6 | F | Mental and behavioural disorders | F10 | Mental and behavioural disorders due to use of alcohol | 0.422305467 | 15 |
| 6 | F | Mental and behavioural disorders | F84 | Pervasive developmental disorders | 0.330254184 | 14 |
| 6 | F | Mental and behavioural disorders | F40 | Phobic anxiety disorders | 0.2858434715 | 13 |
| 7 | O | Pregnancy, childbirth and the puerperium | O26 | Maternal care for other conditions predominantly related to pregnancy | 0.4077711 | 31 |
| 7 | O | Pregnancy, childbirth and the puerperium | O47 | False labour | 0.197829516 | 26 |
| 7 | O | Pregnancy, childbirth and the puerperium | O20 | Haemorrhage in early pregnancy | 0.192177261 | 21 |
| 7 | O | Pregnancy, childbirth and the puerperium | O99 | Other maternal diseases classifiable elsewhere but complicating pregnancy, childbirth and the puerperium | 0.073479532 | 21 |
| 7 | O | Pregnancy, childbirth and the puerperium | O32 | Maternal care for known or suspected malpresentation of fetus | 0.062982469 | 18 |
| 8 | F | Mental and behavioural disorders | F32 | Depressive episode | ######## | 13 |
| 8 | F | Mental and behavioural disorders | F45 | Somatoform disorders | 0.04925552 | 8 |
| 8 | F | Mental and behavioural disorders | F51 | Nonorganic sleep disorders | 0.01291948 | 5 |
| 8 | F | Mental and behavioural disorders | F61 | Mixed and other personality disorders | 0.012112012 | 6 |
| 8 | F | Mental and behavioural disorders | F38 | Other mood [affective] disorders | 0.0113045445 | 5 |
| 9 | F | Mental and behavioural disorders | F90 | Hyperkinetic disorders | 0.6919995395 | 18 |
| 9 | F | Mental and behavioural disorders | F15 | Mental and behavioural disorders due to use of other stimulants, including caffeine | 0.053292855 | 5 |
| 9 | F | Mental and behavioural disorders | F98 | Other behavioural and emotional disorders with onset usually occurring in childhood and adolescence | 0.053292845 | 4 |
| 9 | F | Mental and behavioural disorders | F12 | Mental and behavioural disorders due to use of cannabinoids | 0.040373369 | 5 |
| 9 | F | Mental and behavioural disorders | F44 | Dissociative [conversion] disorders | 0.020186686 | 4 |
| 10 | J | Diseases of the respiratory system | J45 | Asthma | 0.6831176315 | 15 |
| 10 | J | Diseases of the respiratory system | J30 | Vasomotor and allergic rhinitis | 0.49659254 | 9 |
| 10 | J | Diseases of the respiratory system | J98 | Other respiratory disorders | 0.0201866875 | 7 |
| 10 | J | Diseases of the respiratory system | J44 | Other chronic obstructive pulmonary disease | 0.018571751 | 6 |
| 10 | J | Diseases of the respiratory system | J39 | Other diseases of upper respiratory tract | 0.01291948 | 7 |

Table S 9: Dynet top 10 rewired metric to MS vs Control

| name | label | Group | dn | dn_degree | Degree | PageRank | BetweennessCentrality | AverageShortestPathLength | Neighborhood Connectivity |
| --- | --- | --- | --- | --- | --- | --- | --- | --- | --- |
| R2 | Symptoms and signs involving the skin and subcutaneous tissue | Control | 219.29977941782053 | 0.6526779149339896 | 311 | 0.006573 | 0.7887021621380946 | 1.2608695652173914 | 140.91017964071855 |
| R2 | Symptoms and signs involving the skin and subcutaneous tissue | MS | 219.29977941782053 | 0.6526779149339896 | 299 | 0.010226572551605018 | 1.4885970922959286 | 1.2626262626262625 | 98.67682926829268 |
| G9 | Other disorders of the nervous system | Control | 159.31367466645065 | 0.6372546986658026 | 199 | 0.00414 | 0.050677 | 1.4927536231884058 | 152.66379310344828 |
| G9 | Other disorders of the nervous system | MS | 159.31367466645065 | 0.6372546986658026 | 188 | 0.006684 | 0.23204015579513046 | 1.5808080808080809 | 113.6875 |
| H5 | Disorders of ocular muscles, binocular movement, accommodation and refraction | Control | 150.48042545204433 | 0.45189316952565867 | 320 | 0.006855 | 0.6471829314123635 | 1.2318840579710144 | 138.85294117647058 |
| H5 | Disorders of ocular muscles, binocular movement, accommodation and refraction | MS | 150.48042545204433 | 0.45189316952565867 | 261 | 0.008809 | 0.6836940219677294 | 1.3737373737373737 | 106.15972222222223 |
| Z9 | Persons with potential health hazards related to family and personal history and certain conditions influencing health status | Control | 134.4815703668351 | 0.39207454917444634 | 337 | 0.007117 | 0.5750602002814145 | 1.2028985507246377 | 136.56497175141243 |
| Z9 | Persons with potential health hazards related to family and personal history and certain conditions influencing health status | MS | 134.4815703668351 | 0.39207454917444634 | 236 | 0.007553 | 0.7422317060227637 | 1.398989898989899 | 106.92424242424242 |
| M0 | Arthropathies: Infectious arthropathies | Control | 133.49819618419036 | 0.4184896432106281 | 312 | 0.006624 | 0.4210415528411959 | 1.2608695652173914 | 140.35329341317365 |
| M0 | Arthropathies: Infectious arthropathies | MS | 133.49819618419036 | 0.4184896432106281 | 181 | 0.006243 | 0.176351 | 1.5707070707070707 | 117.59433962264151 |
| N1 | Renal tubulo-interstitial diseases | Control | 130.87329727801256 | 0.46409 | 276 | 0.005862 | 0.21383211269602298 | 1.3478260869565217 | 143.82119205298014 |
| N1 | Renal tubulo-interstitial diseases | MS | 130.87329727801256 | 0.46409 | 100 | 0.003544 | 0.068843 | 1.7525252525252526 | 119.35820895522389 |
| M3 | Systemic connective tissue disorders | Control | 129.94659996959524 | 0.4191825805470814 | 297 | 0.006211 | 0.23875443833608695 | 1.289855072463768 | 145.9933774834437 |
| M3 | Systemic connective tissue disorders | MS | 129.94659996959524 | 0.4191825805470814 | 174 | 0.006007 | 0.43017840917049754 | 1.5858585858585859 | 113.16822429906541 |
| E8 | Metabolic disorders | Control | 128.32934693426117 | 0.5052336493474849 | 238 | 0.005308 | 0.10411452849453617 | 1.463768115942029 | 147.34558823529412 |
| E8 | Metabolic disorders | MS | 128.32934693426117 | 0.5052336493474849 | 94 | 0.003446 | 0.079594 | 1.792929292929293 | 120.11475409836065 |
| J1 | Influenza and pneumonia | Control | 127.53467489273454 | 0.4490657566645582 | 264 | 0.005516 | 0.18627538235935256 | 1.357487922705314 | 144.5974025974026 |
| J1 | Influenza and pneumonia | MS | 127.53467489273454 | 0.4490657566645582 | 147 | 0.005646 | 0.4509133167146936 | 1.6565656565656566 | 114.52 |
| R4 | Symptoms and signs involving cognition, perception, emotional state and behaviour | Control | 126.27377556148191 | 0.388535 | 310 | 0.00666 | 0.30592702781273307 | 1.2801932367149758 | 141.74096385542168 |
| R4 | Symptoms and signs involving cognition, perception, emotional state and behaviour | MS | 126.27377556148191 | 0.388535 | 244 | 0.00836 | 0.5915331779568456 | 1.4191919191919191 | 106.38571428571429 |

Figure S 1: Top 10 Disease Clusters for MS Groups. Network-based clustering was employed to identify tightly interconnected diagnostic communities, offering insights into co-occurring diseases and patient trajectories preceding MS onset. To identify diagnostically meaningful subgroups, we applied the MCL to both the MS and non-MS networks.

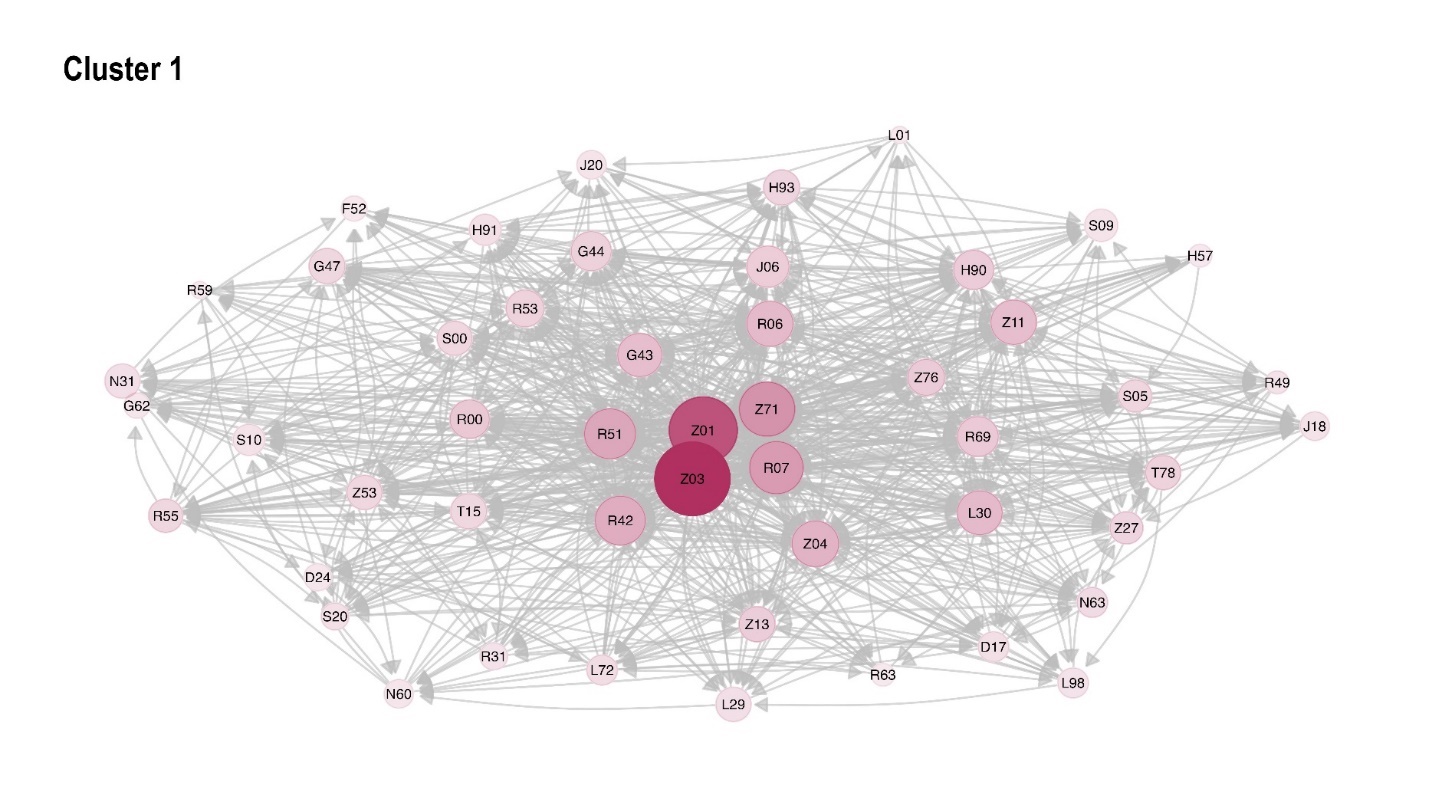

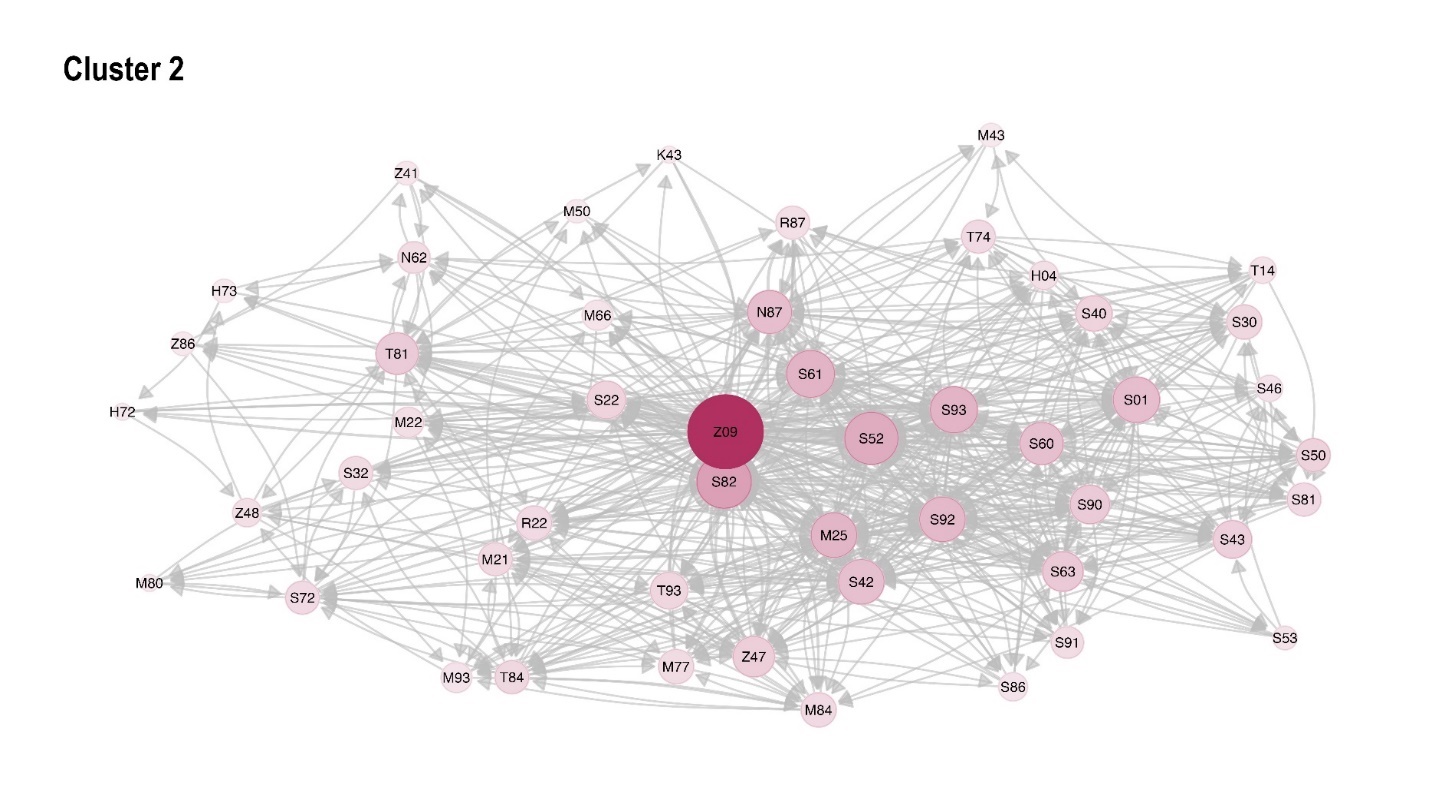

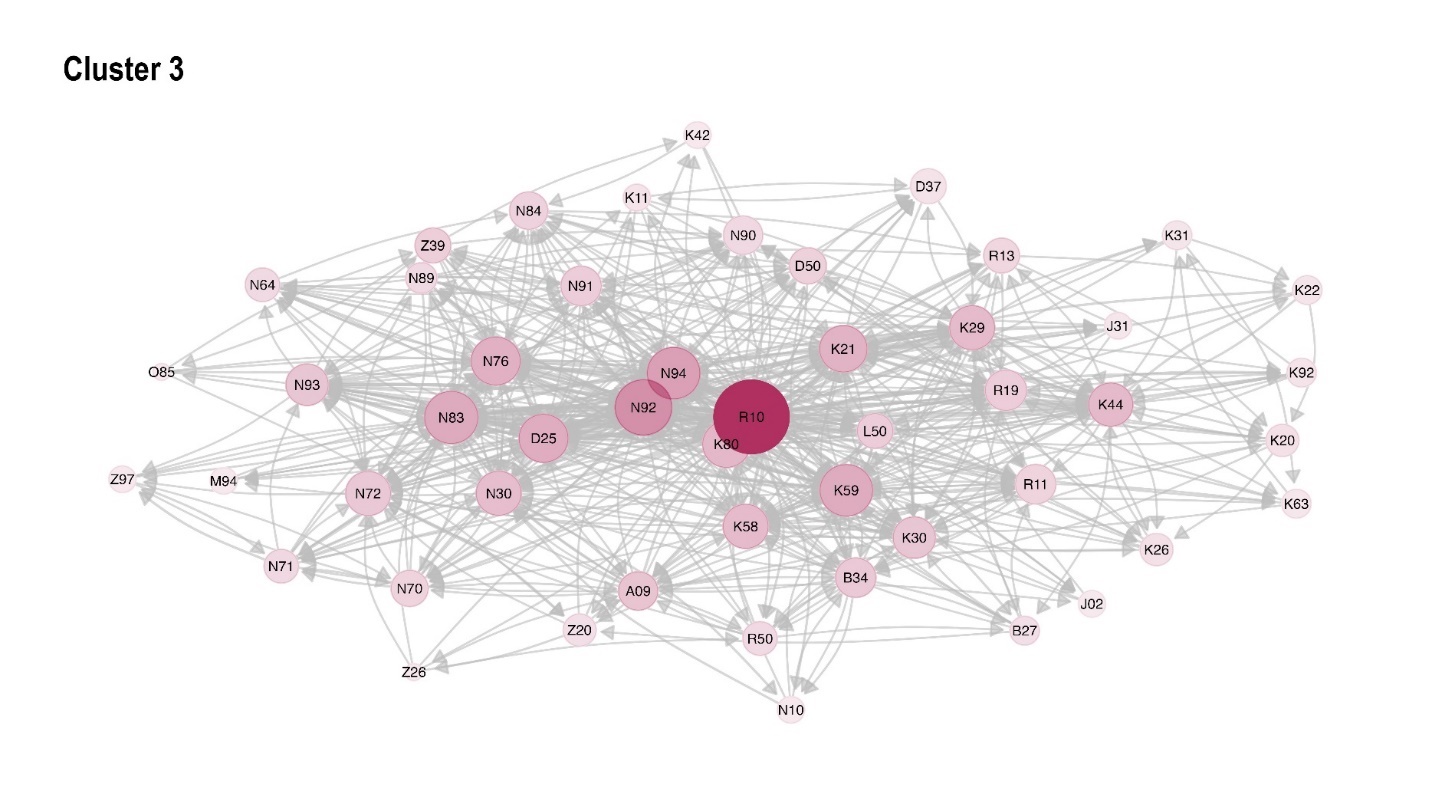

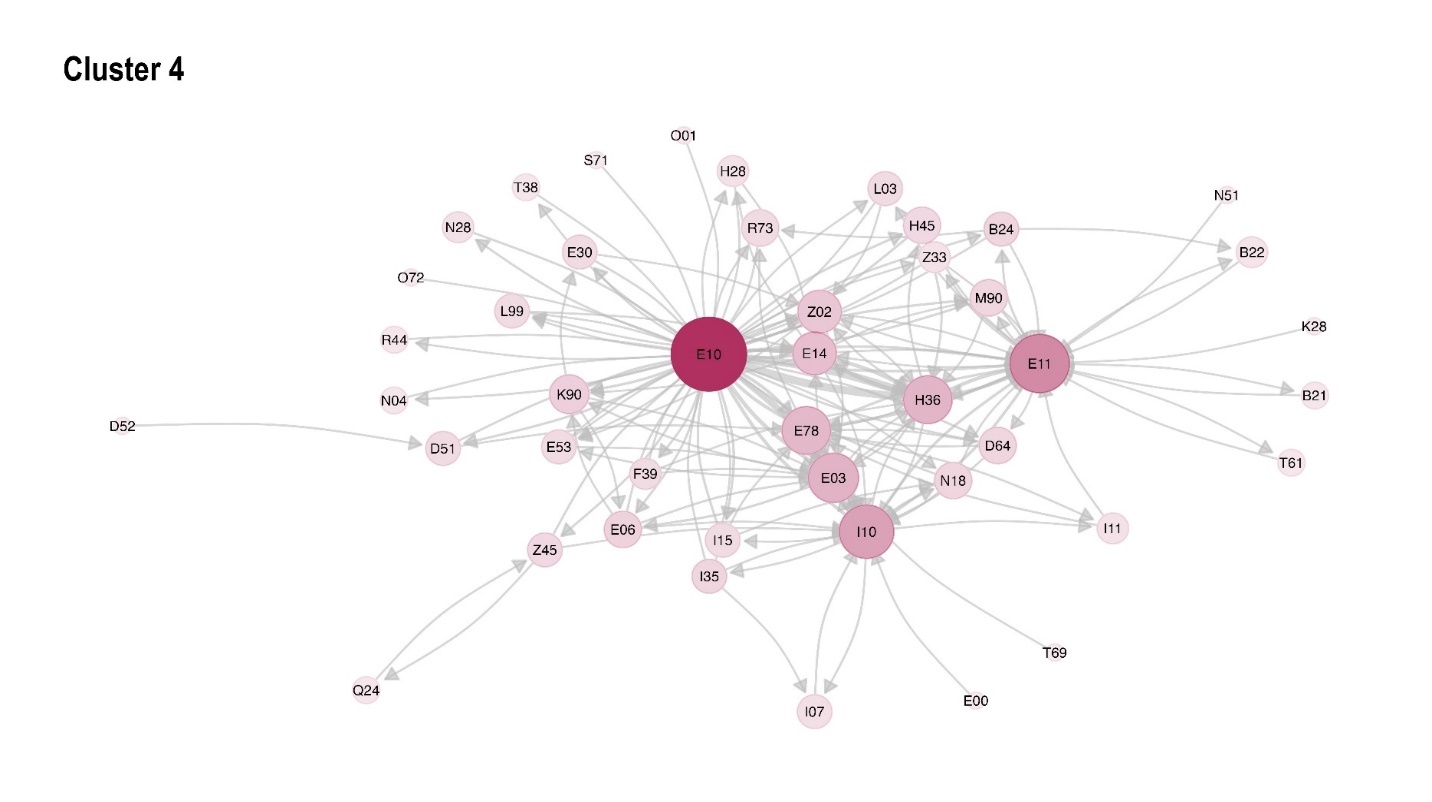

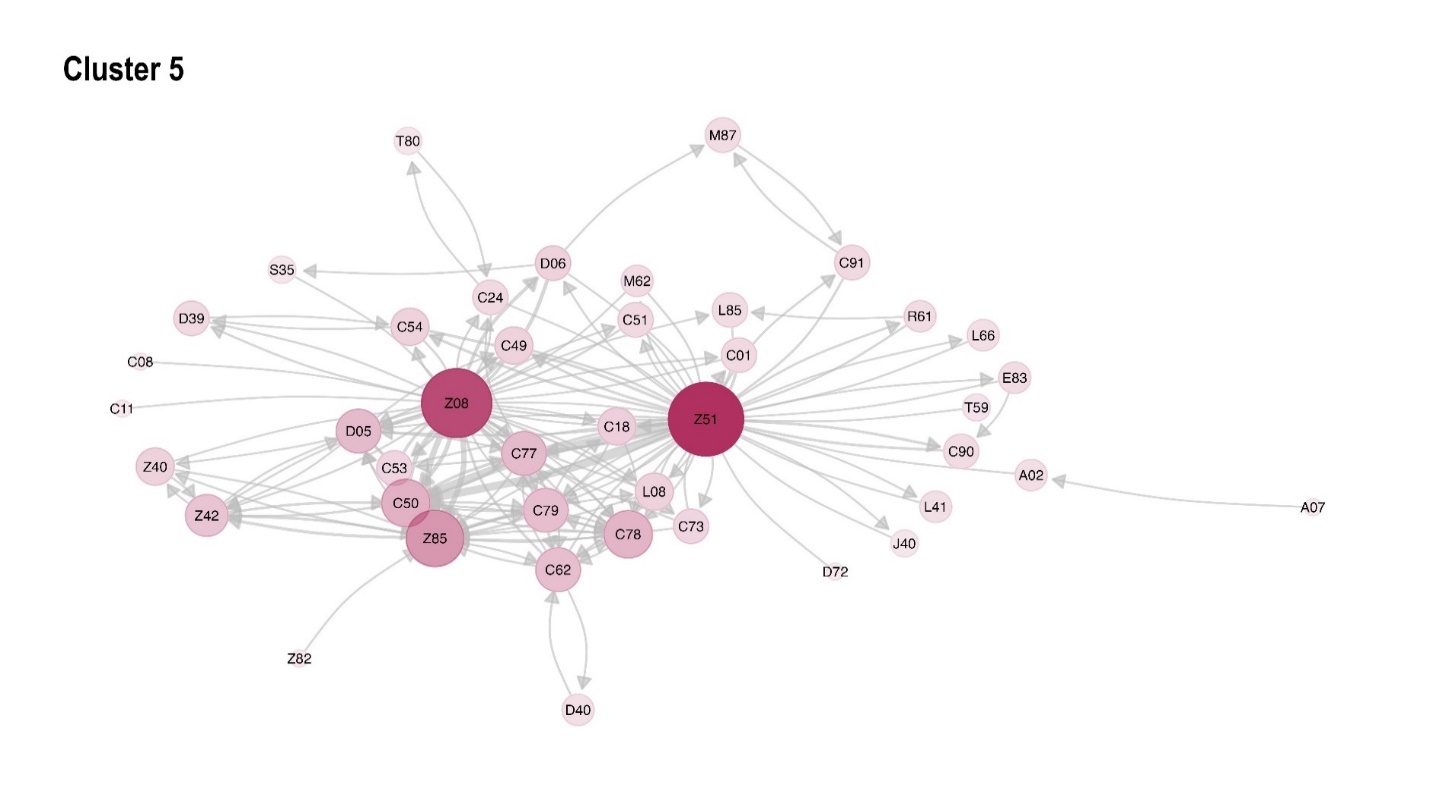

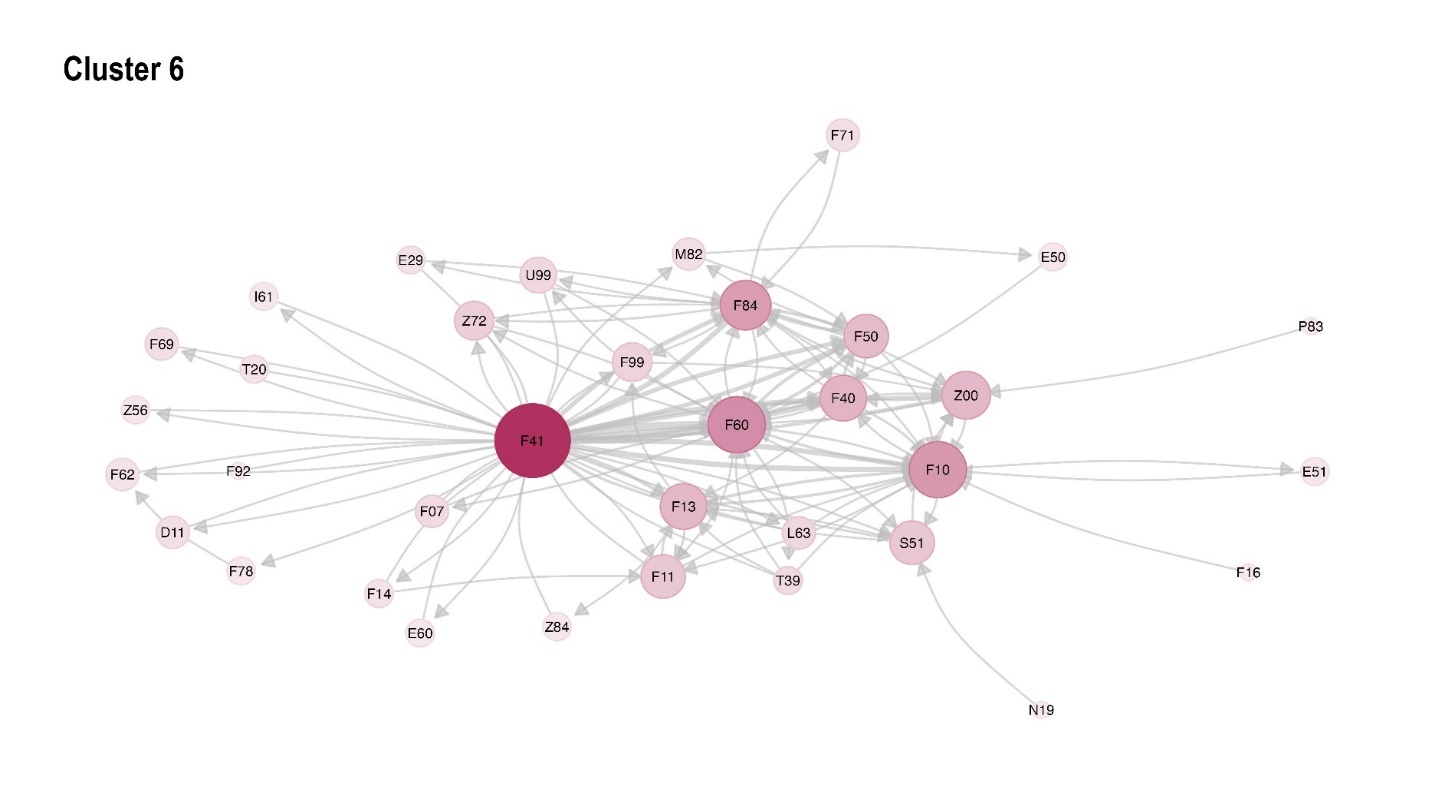

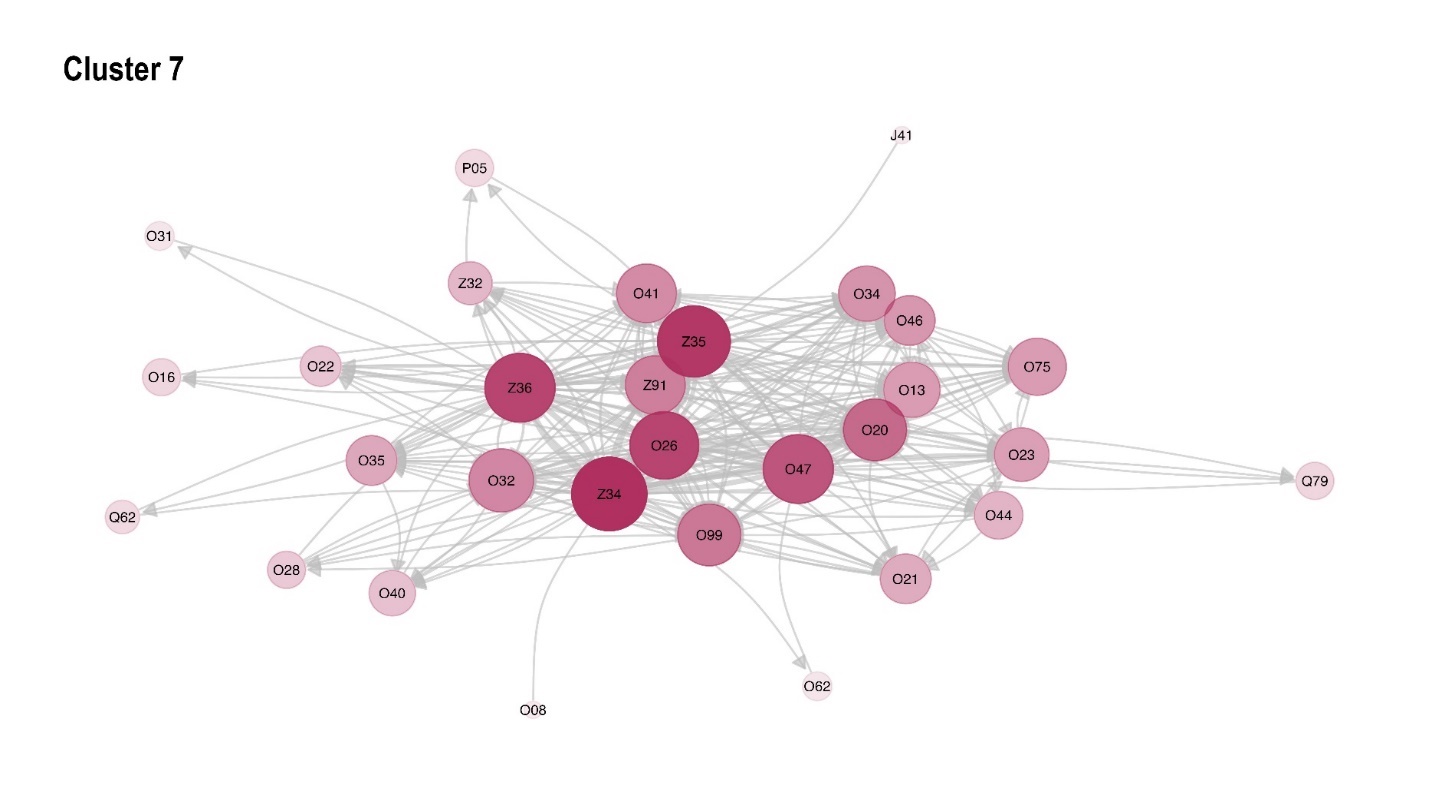

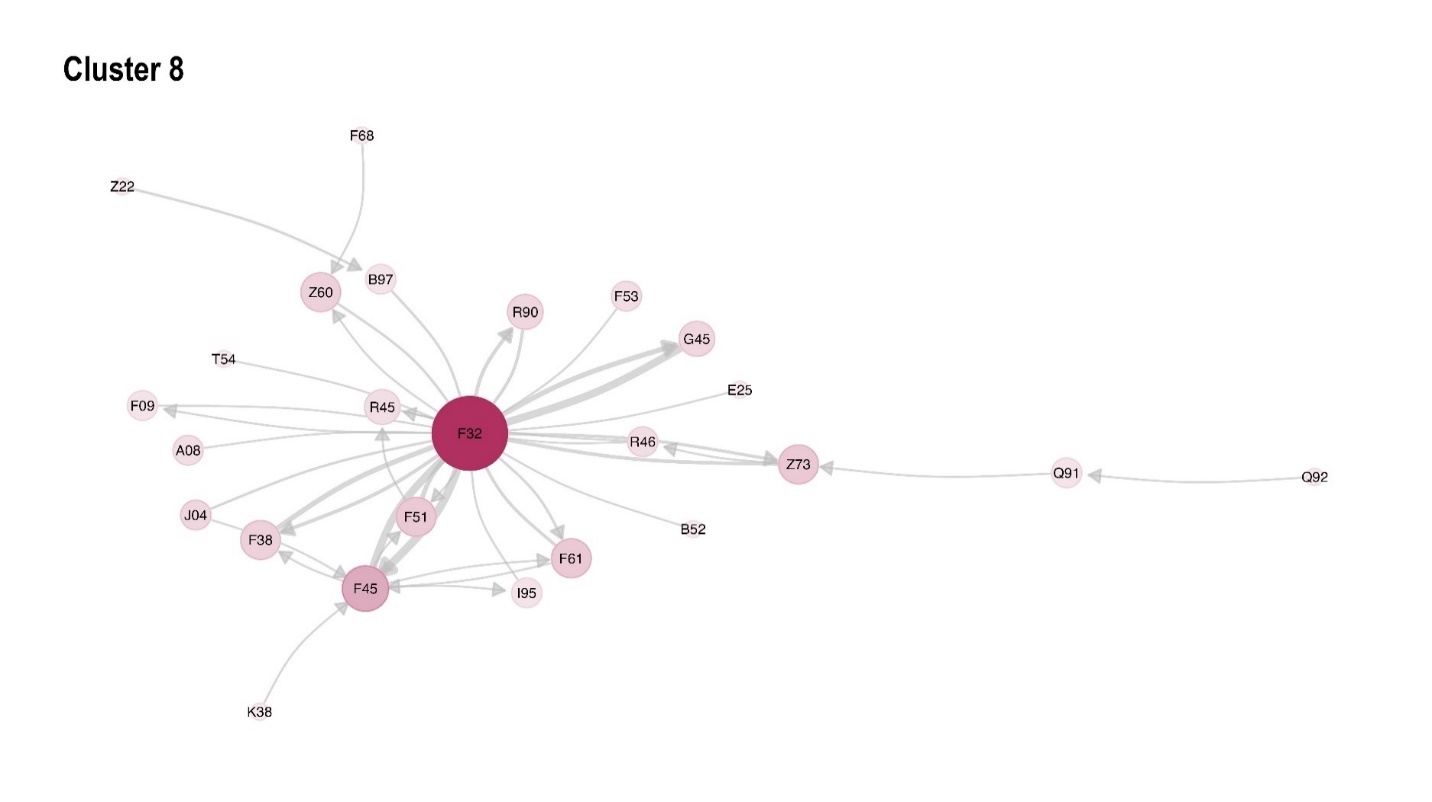

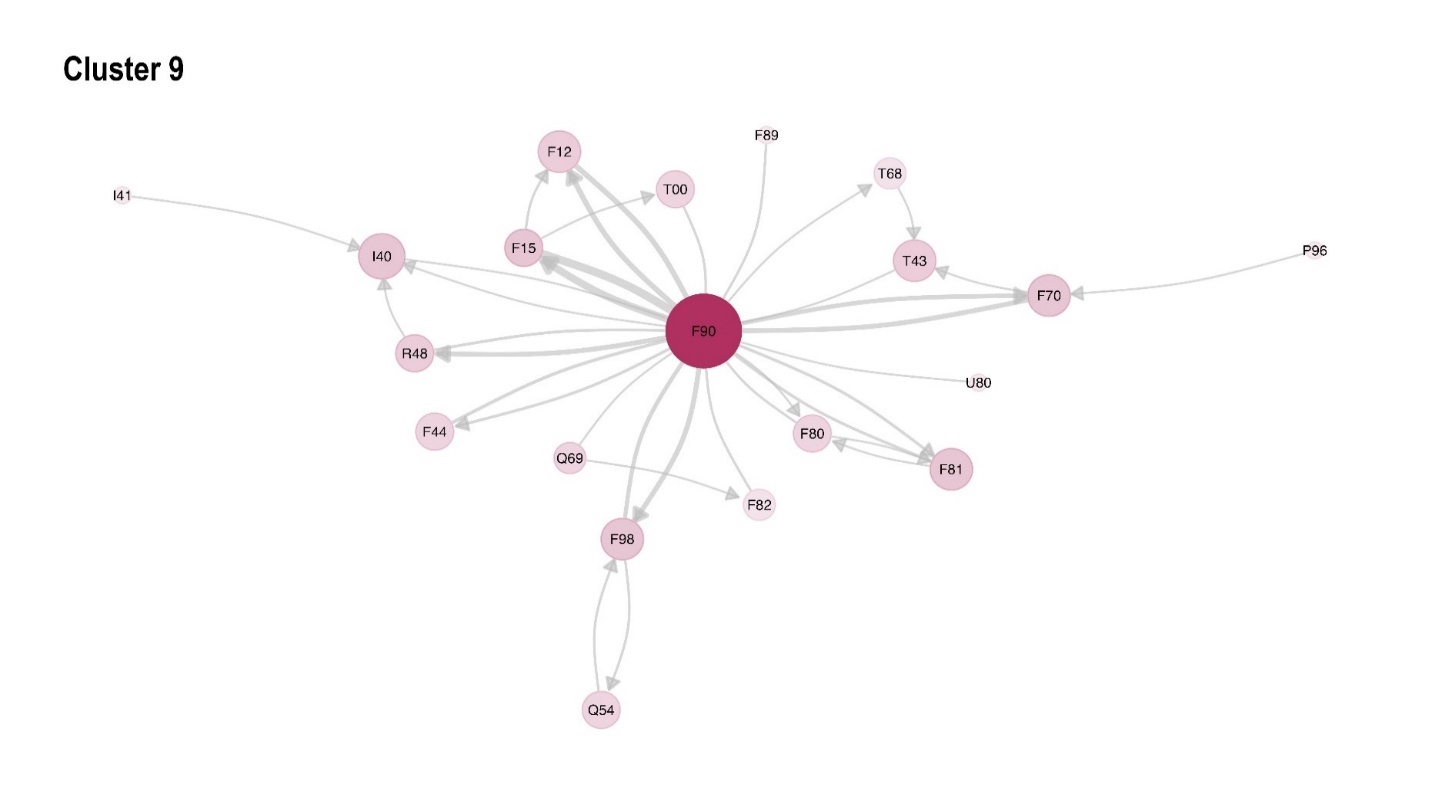

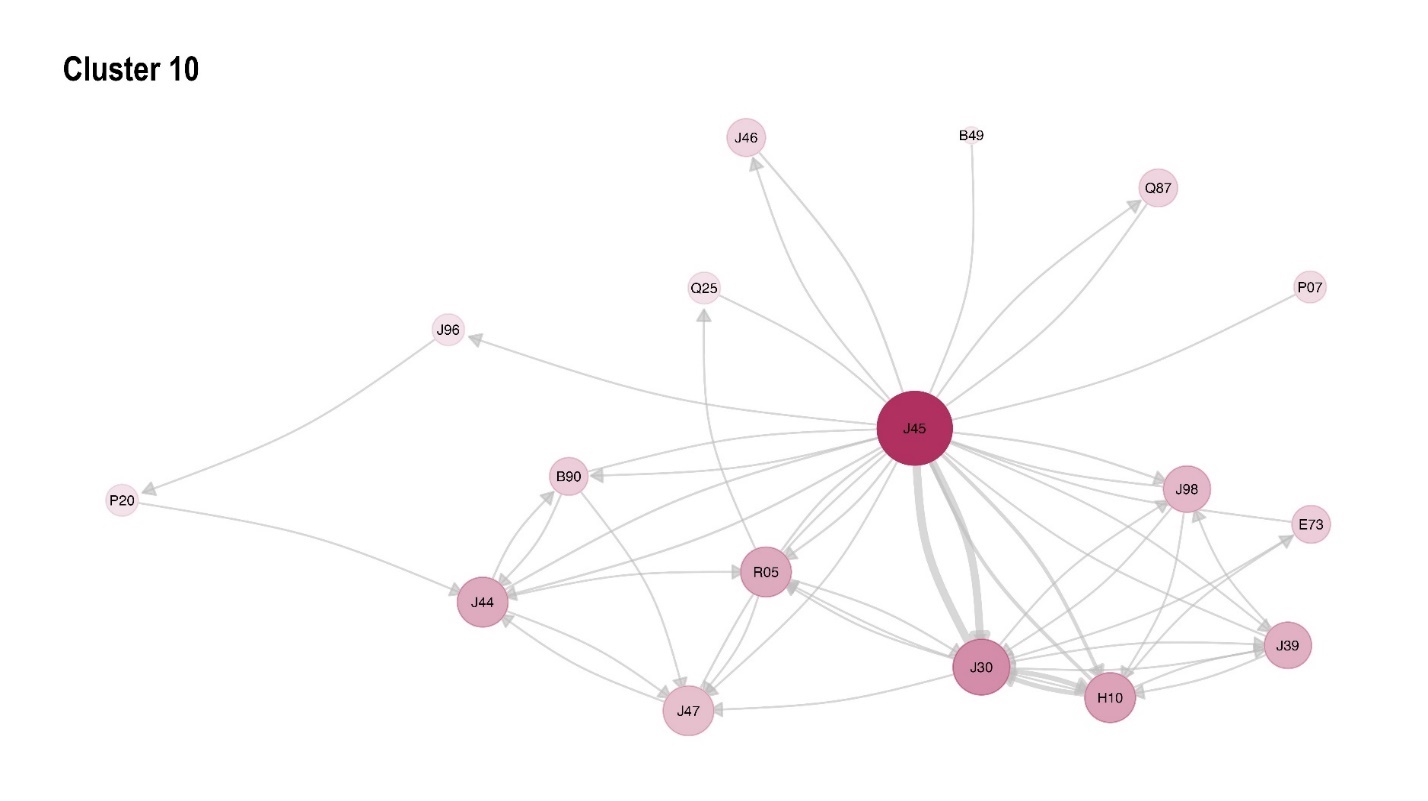

Figure S 2: Top 10 Disease Clusters for Controls. Network-based clustering was employed to identify tightly interconnected diagnostic communities, offering insights into co-occurring diseases and patient trajectories preceding MS onset. To identify diagnostically meaningful subgroups, we applied the MCL to both the MS and non-MS networks.

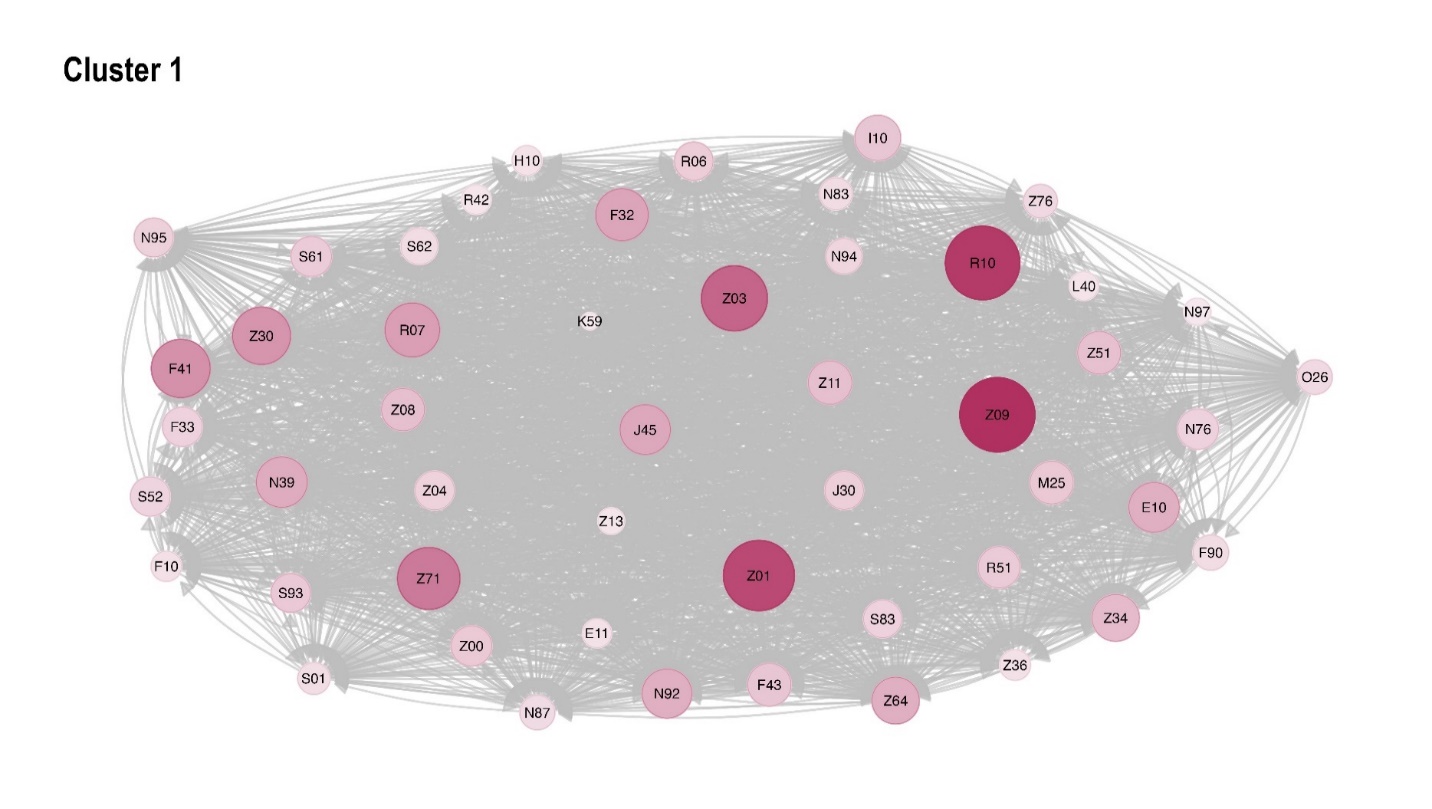

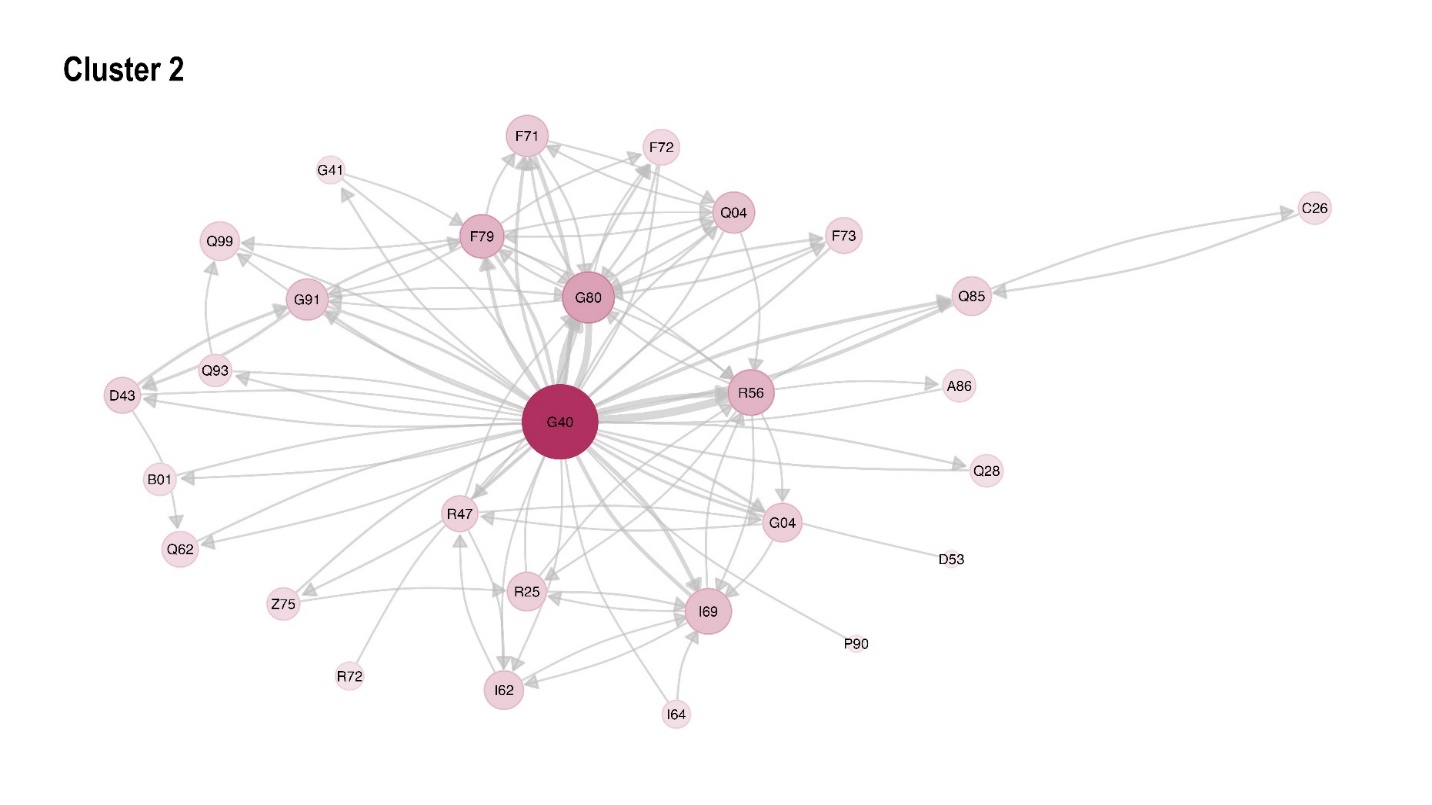

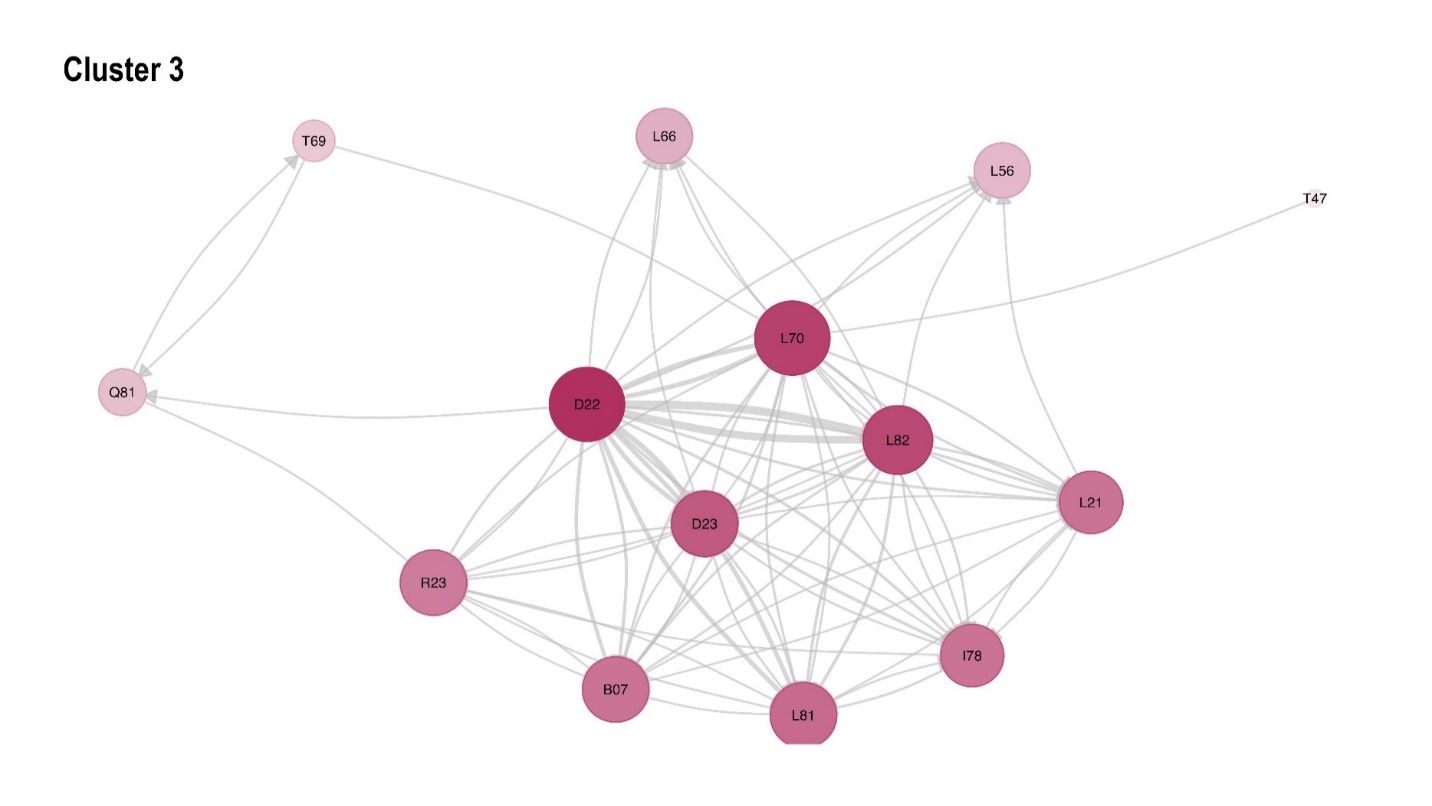

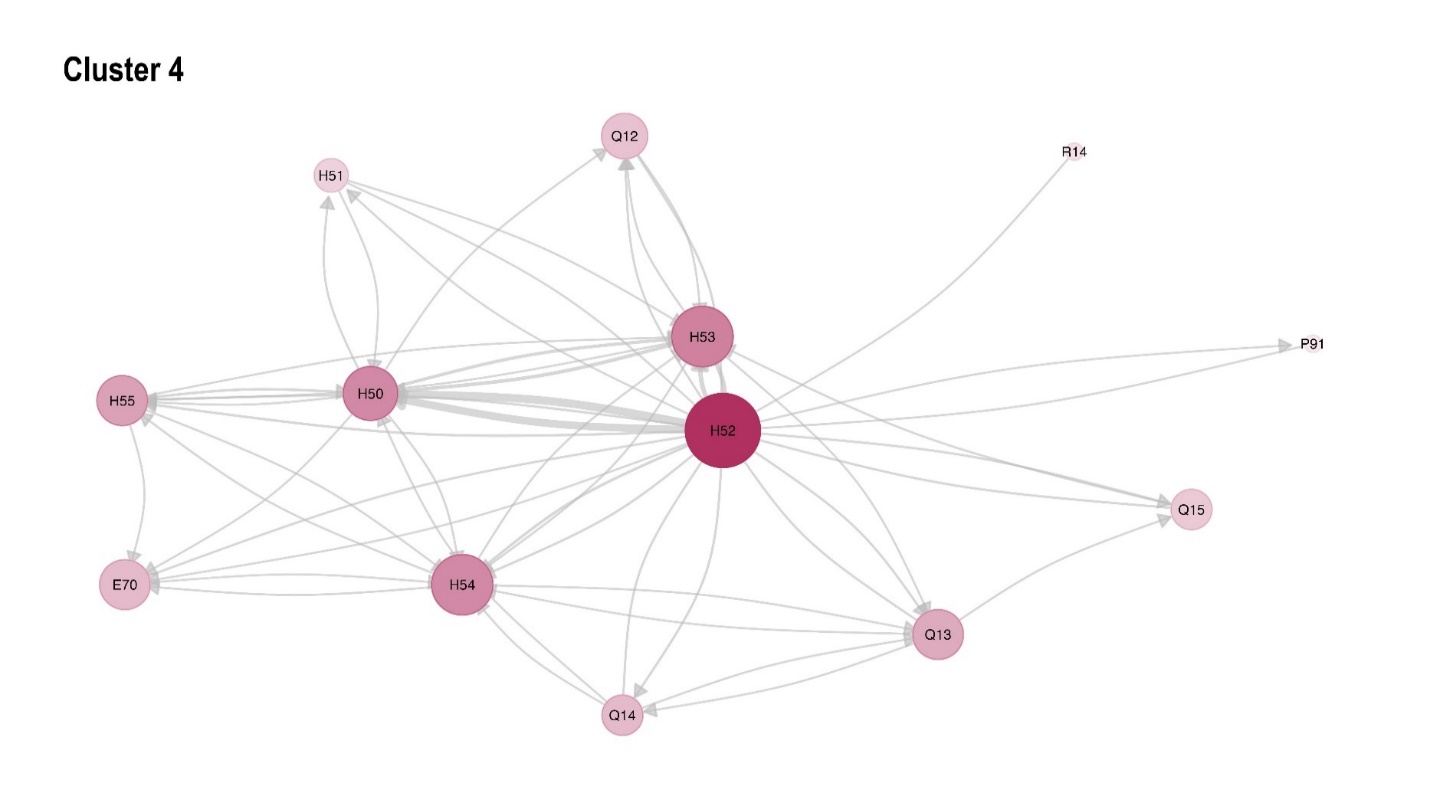

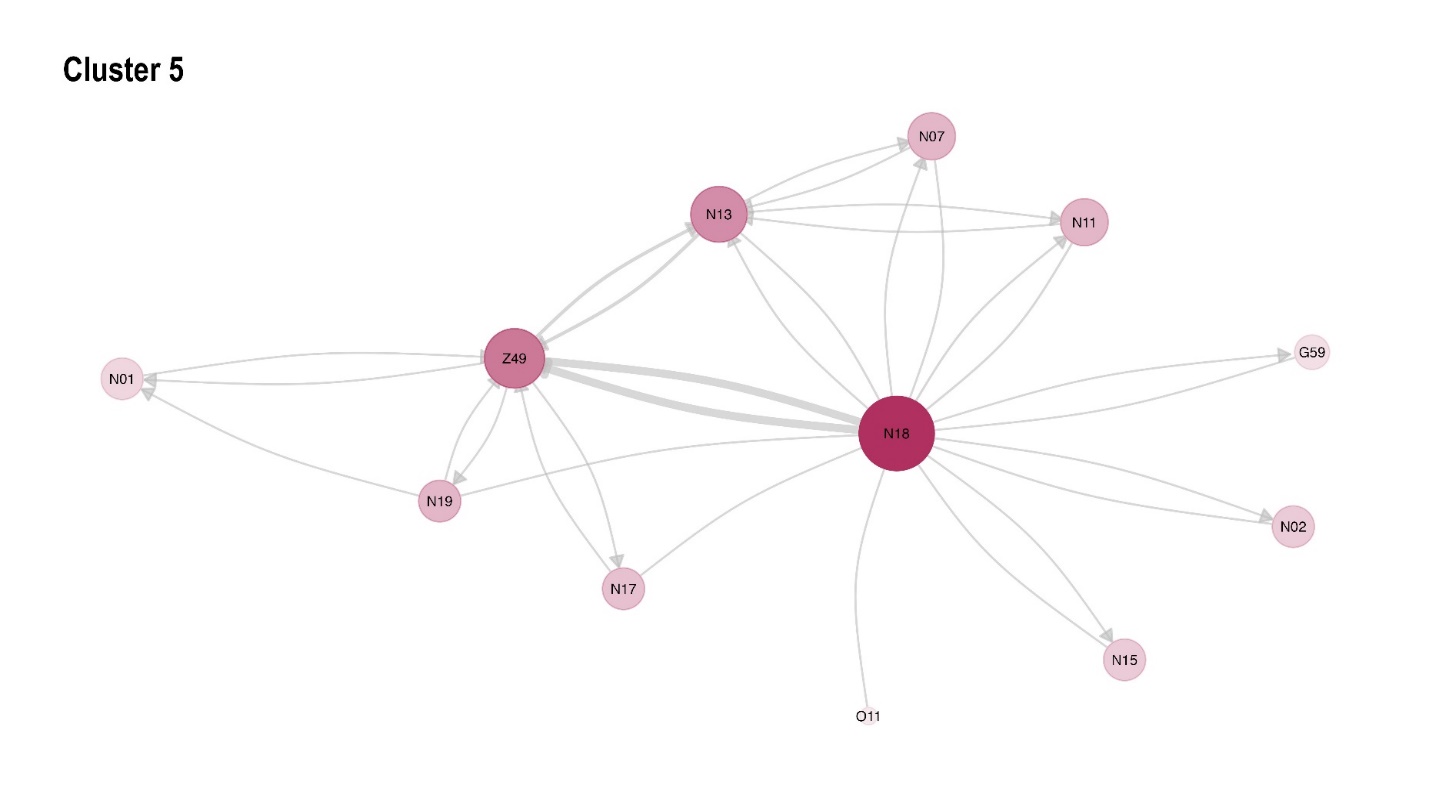

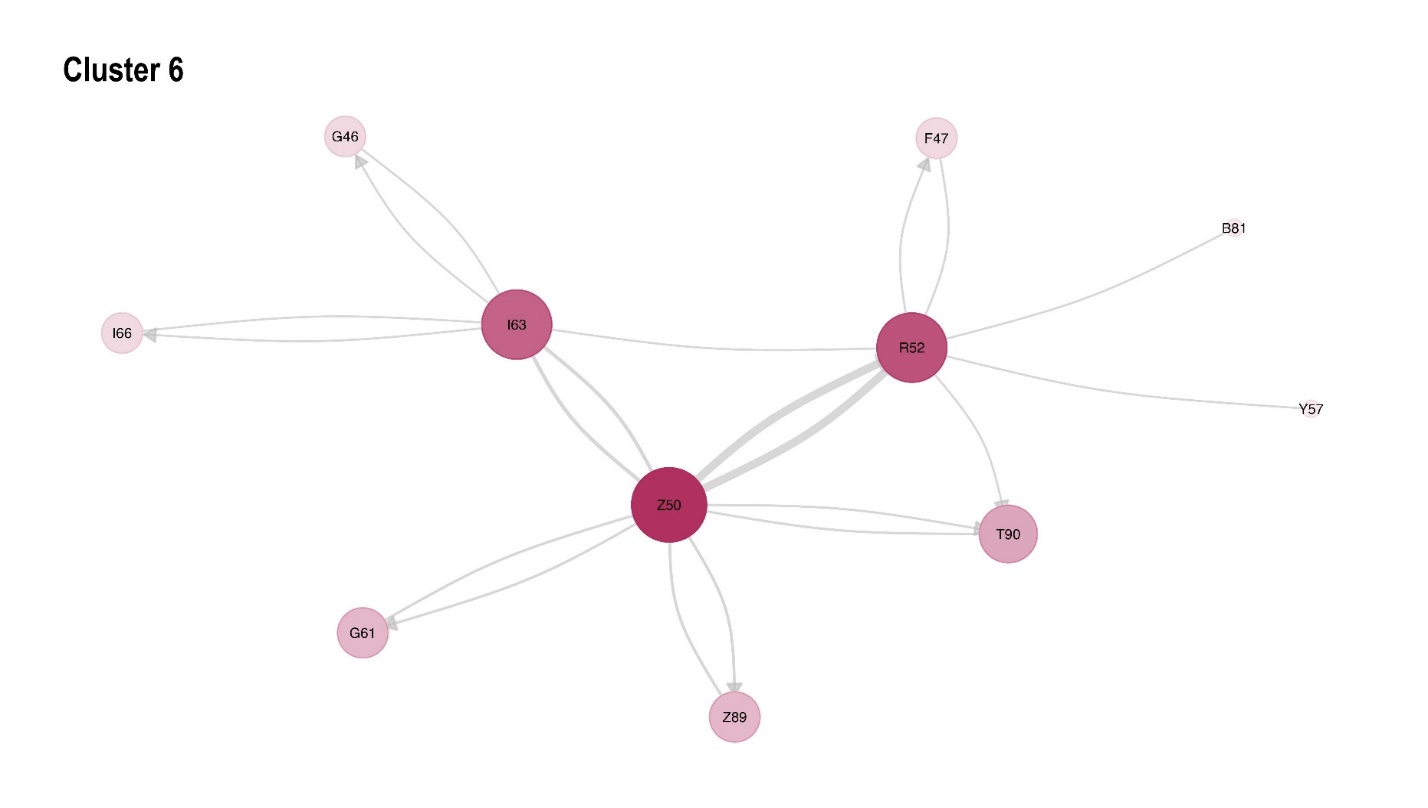

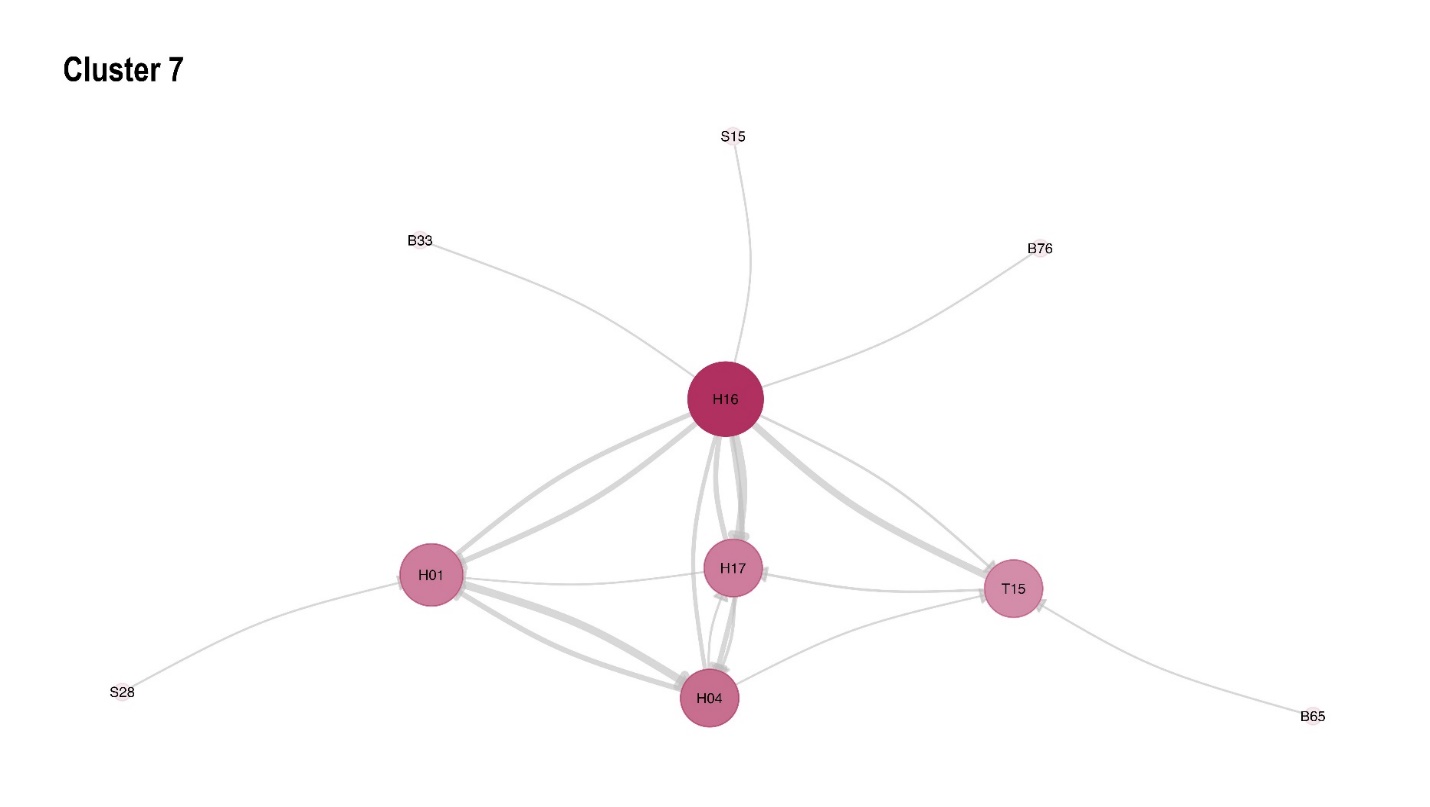

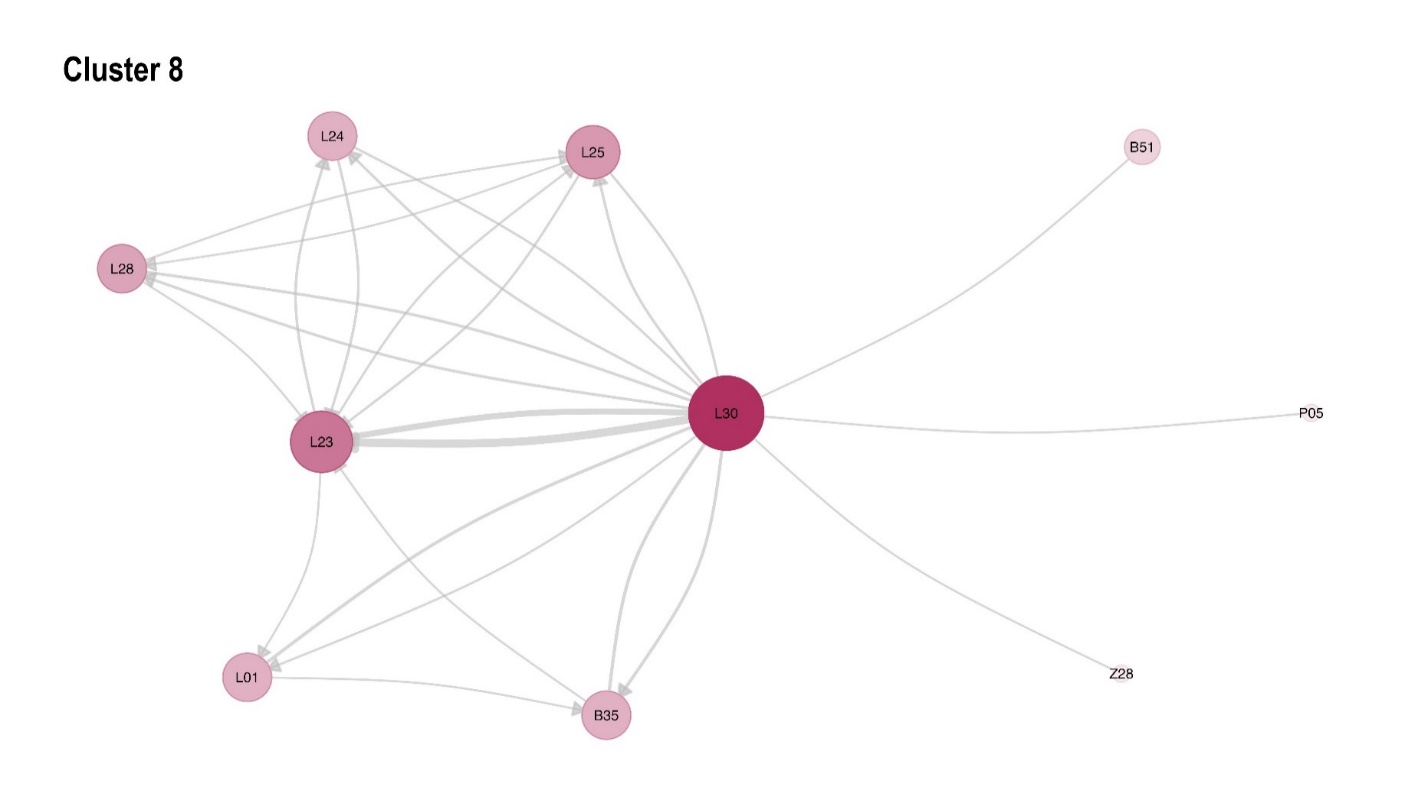

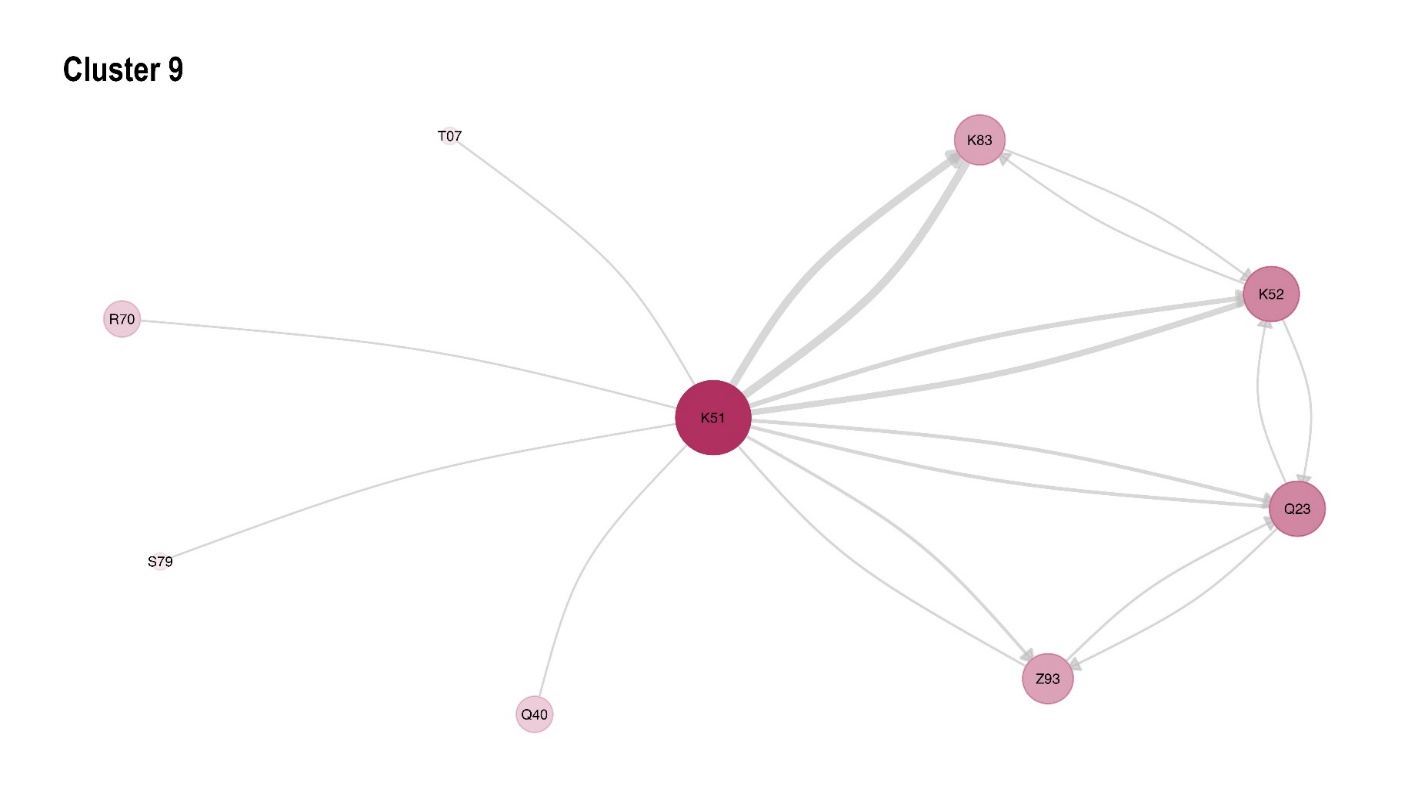

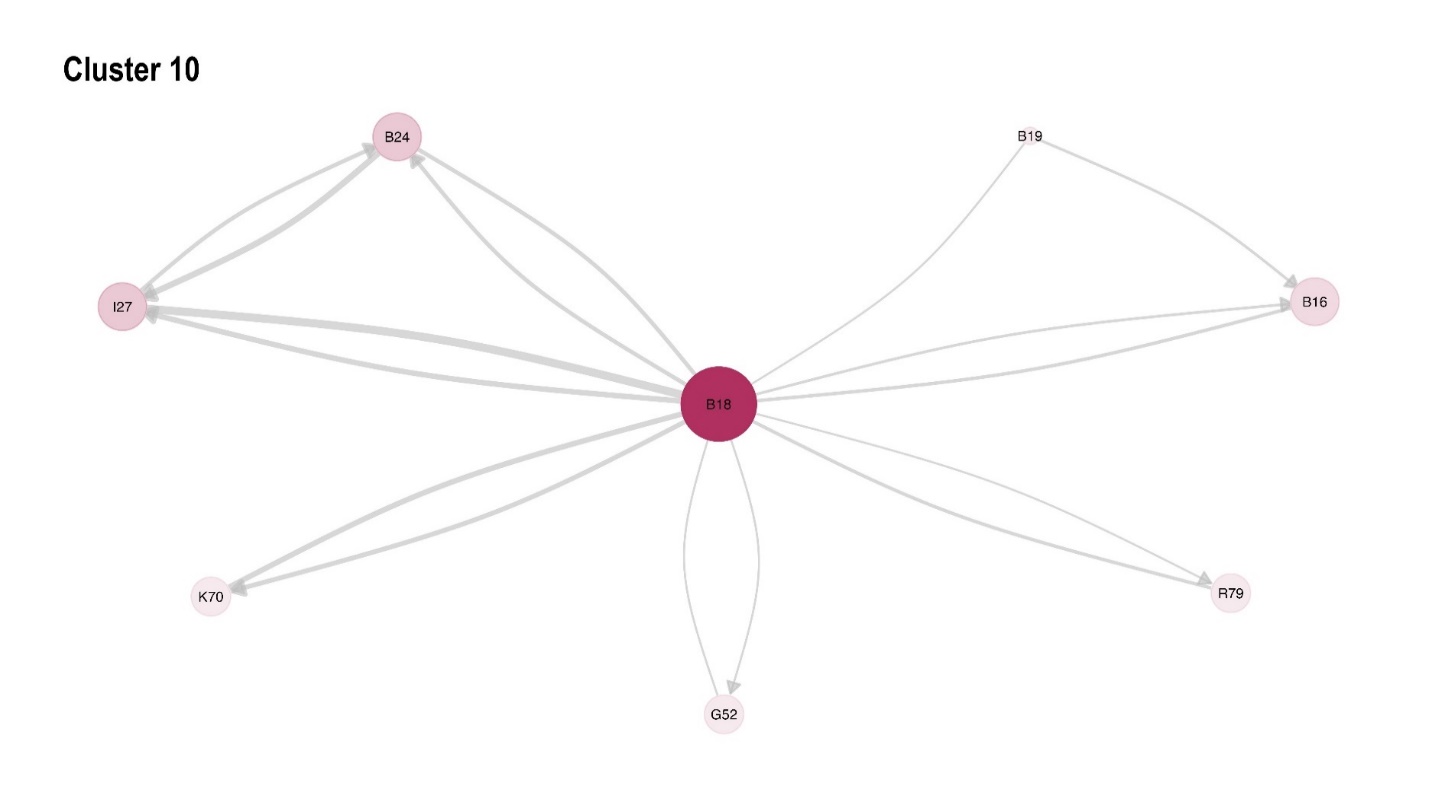

Figure S 3: Comparison between dominant ICD chapters

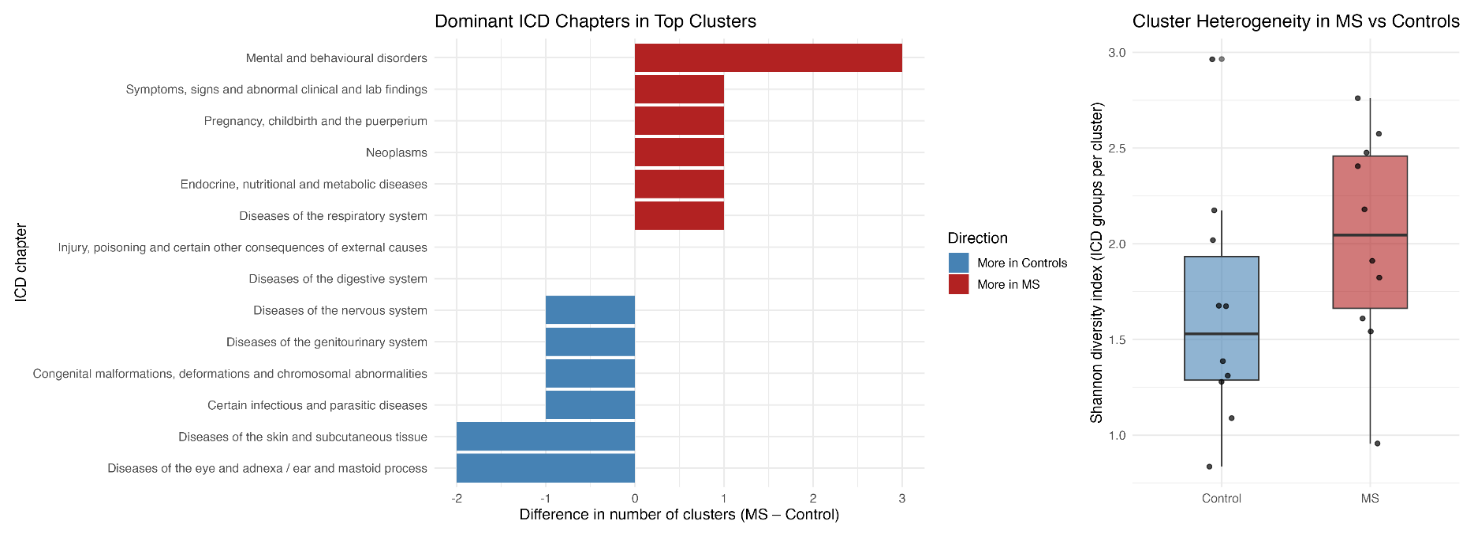
